# Who Reaches Surgery, Who Survives? Skilled Intrapartum Care and Early Neonatal Death in a National Cohort of Nigerian Facility Births

**DOI:** 10.1101/2025.11.24.25340930

**Authors:** Sunday A. Adetunji, Bukola Adetunji

## Abstract

**Background:** Nigeria accounts for one of the world’s largest burdens of neonatal deaths despite rising coverage of facility delivery and skilled birth attendance (SBA). How intrapartum care is allocated within facilities—and what it achieves for early neonatal survival—remains poorly understood.

**Methods:** We analyzed nationally representative data from the 2018 Nigeria Demographic and Health Survey, restricting to the most recent singleton birth in the preceding 5 years that occurred in a health facility. Exposures were SBA (doctor, nurse, or midwife vs none) and caesarean versus vaginal delivery. The primary outcome was early neonatal death (0–6 days); secondary outcome was maternal report of “very small” size at birth. We used survey-weighted descriptive statistics to characterize socio-economic and obstetric gradients in care, and propensity-score overlap weighting combined with the DHS survey design to estimate absolute and relative associations between intrapartum care and outcomes among women in the region of covariate overlap. Pre-specified subgroup, equity, and scenario analyses explored effect modification and potentially avoidable deaths.

**Results:** The analytic cohort comprised 5,860 facility-based singleton births, representing approximately 5.7 million Nigerian facility births. SBA coverage was 96.6% and caesarean delivery accounted for 12.1% of births; both were strongly concentrated among urban, wealthier, and better-educated women. The survey-weighted prevalence of early neonatal death was 3.8% and of very small size at birth 2.7%. In crude analyses, early neonatal mortality was higher among births with SBA than without (risk difference, 2.2 percentage points; 95% CI −0.1 to 4.4), reflecting strong confounding by indication. After overlap weighting, the risk difference comparing SBA with non-SBA was 2.5 percentage points (95% CI 0.2 to 4.8; risk ratio 2.59, 95% CI 0.70 to 9.62). For caesarean versus vaginal delivery, adjusted risk of early neonatal death was 3.9% versus 3.3% (risk difference 0.6 percentage points; 95% CI −1.1 to 2.4; risk ratio 1.20, 95% CI 0.75 to 1.90), ruling out large harmful effects but providing no strong evidence of benefit. Associations with very small size at birth were small and imprecise for both exposures. Joint-pattern analyses showed the lowest mortality for skilled vaginal births and higher risks for both non-skilled vaginal and skilled caesarean births, against a background of steep wealth, geographic, and facility-sector gradients. Scenario models suggested that extending competent SBA and timely, indicated caesarean delivery to poor and rural women could prevent a substantial share of facility-based early neonatal deaths.

**Conclusions:** In Nigeria, intrapartum expertise and surgical capacity are scarce and inequitably distributed, yet medically indicated caesarean delivery does not appear to confer large additional survival benefit within current system constraints. Early neonatal deaths cluster where SBA and caesarean access are lowest and quality gaps are greatest. Closing Nigeria’s neonatal survival gap will require not only higher coverage of facility births, but also reliable, equitable access to timely skilled intrapartum care and emergency obstetric services of adequate quality.

## 1. INTRODUCTION

Each year, an estimated 2.3 million newborns die in the first 28 days of life, and nearly half of all under-five deaths now occur during this brief neonatal period (United Nations Inter-agency Group for Child Mortality Estimation [UN IGME], 2022; UNICEF, 2023). Complications of preterm birth, intrapartum-related events (including birth asphyxia), and severe neonatal infections account for most of these deaths, with the highest risks concentrated in sub-Saharan Africa and South Asia (Lawn et al., 2014; Wang et al., 2016). Although under-five mortality has fallen substantially since 1990, progress for newborns has been slower, and the global target of no more than 10 neonatal deaths per 1,000 live births remains out of reach for many high-burden countries (Lawn et al., 2014; UN IGME, 2022).

Nigeria sits at the epicenter of this unfinished agenda. National survey and modelled estimates indicate a neonatal mortality rate of about 39 deaths per 1,000 live births in 2018, among the highest in the world and more than double the recent global average (National Population Commission [NPC] & ICF, 2019; Kunnuji et al., 2022). Nigeria contributes one of the largest absolute numbers of neonatal deaths worldwide; even modest improvements in early neonatal survival in this single country would therefore translate into large global gains (UNICEF, 2023). Recent analyses of Demographic and Health Surveys (DHS) and Multiple Indicator Cluster Surveys suggest that while neonatal mortality in Nigeria has declined compared with earlier decades, progress has plateaued and in some regions reversed, with persistent inequalities by geography, household wealth, and maternal education (Kunnuji et al., 2022; Nwanze et al., 2023; Wammanda et al., 2022).

Over the same period, the place and type of childbirth care in low- and middle-income countries have shifted markedly. Coverage of facility delivery and skilled birth attendance (SBA) has risen, including in Nigeria, where a growing share of births now occur in health facilities with a doctor, nurse, or midwife present (NPC & ICF, 2019; UNICEF, 2023). Yet declines in maternal and neonatal mortality have not kept pace with this expansion of contact coverage. This disconnect has crystallized a consensus that high-quality, timely intrapartum and immediate postnatal care—not facility contact alone—is essential to avert preventable deaths (Koblinsky et al., 2016; Kruk et al., 2016; World Health Organization [WHO], 2016). The WHO quality-of-care framework for maternal and newborn health explicitly emphasizes competent intrapartum care, effective referral and emergency obstetric services, and continuous monitoring around the time of birth as the critical levers for improving outcomes (WHO, 2016).

Within this framework, two intrapartum modalities are especially salient for neonatal survival: the presence of a skilled birth attendant and access to timely caesarean section when clinically indicated. SBA is a core global indicator and is assumed to capture both technical competence and the capacity to respond rapidly to complications. Caesarean delivery, in turn, is a lifesaving procedure for obstructed labour, fetal distress, malpresentation, and selected maternal indications, but it carries appreciable risk when performed inappropriately or in settings with limited perioperative and neonatal care capacity (Sobhy et al., 2019; WHO, 2015). The Lancet Maternal Health Series framed this duality as a spectrum from “too little, too late” to “too much, too soon,” underscoring that both underuse and overuse of obstetric interventions can be harmful, particularly where health systems are weak (Miller et al., 2016; Koblinsky et al., 2016).

Nigeria exemplifies this tension. Nationally, overall caesarean section rates remain well below the 10–15% range often considered compatible with meeting obstetric need, with earlier DHS analyses suggesting that only around 2–3% of live births are delivered by caesarean section (Adewuyi et al., 2019). Within facilities and among wealthier, urban, and more educated women, however, caesarean use rises sharply, whereas poor, rural, and less educated women face substantial barriers to both SBA and surgical delivery (Adewuyi et al., 2019; Okyere et al., 2022; Olukade et al., 2022). Hospital and program evaluations from Nigeria document high burdens of intrapartum complications, severe neonatal morbidity, and early neonatal deaths despite facility birth, pointing to serious gaps in timeliness, monitoring, and the organization of emergency obstetric and newborn care (Tukur et al., 2022; Audu et al., 2021; Okonofua et al., 2020, 2023). These patterns suggest that lifesaving intrapartum expertise and surgical capacity are not only scarce but also rationed along socio-economic and geographic lines.

Despite a growing body of work, several critical gaps remain. First, most national analyses of neonatal mortality in Nigeria treat mode of delivery and SBA as background covariates or focus on population-level determinants, rather than interrogating how intrapartum care is allocated among women who reach facilities and how that allocation shapes early neonatal outcomes (Kunnuji et al., 2022; Wammanda et al., 2022). Second, much of the evidence on caesarean outcomes in low- and middle-income settings comes from single hospitals or referral networks, limiting generalizability and often lacking robust adjustment for socio-economic and obstetric confounding (Sobhy et al., 2019; Tukur et al., 2022). Third, we know relatively little about how SBA and caesarean delivery perform in combination within real health systems—for example, whether the highest-risk fetuses are successfully triaged into skilled surgical pathways, or whether survival benefits are attenuated by delays and quality deficits along the intrapartum continuum (Kruk et al., 2016; WHO, 2016; Okonofua et al., 2023). Finally, few nationally representative studies have applied modern causal inference methods to estimate the absolute and relative effects of SBA and caesarean delivery on early neonatal death within the subset of births that already occur in facilities, where quality-improvement interventions are most directly actionable.

The 2018 Nigeria DHS offers a unique opportunity to address these gaps. It provides nationally representative data on place of delivery, type of provider, mode of birth, and early neonatal survival, alongside detailed information on maternal socio-demographic characteristics, antenatal care, and household wealth (NPC & ICF, 2019). Because the survey enumerates both facility and non-facility births, it allows us to focus on the intrapartum “last mile” among women who reach a facility while still situating those births within the broader equity landscape. Coupled with advances in propensity-score overlap weighting and survey-weighted causal estimation, these data permit a move beyond descriptive coverage statistics toward policy-relevant estimates of how intrapartum care is distributed and what it achieves for newborn survival (Li et al., 2018; Austin, 2021).

In this study, we restrict attention to singleton births that occurred in health facilities in the five years preceding the 2018 survey and pursue four linked aims. First, we describe the distribution of obstetric and socio-economic risk across key intrapartum modalities—skilled versus non-skilled attendance and caesarean versus vaginal delivery—to illuminate how Nigerian facilities allocate intrapartum care. Second, we estimate the association of SBA with early neonatal death (0–7 days) among facility-based singleton births, using survey-weighted propensity-score overlap weighting to approximate causal contrasts between women who did and did not receive SBA but were otherwise comparable on measured risk factors. Third, we undertake an analogous analysis for caesarean versus vaginal delivery, focusing on women whose clinical profiles make either mode of birth plausible, and assess whether these associations differ across obstetric risk strata, including parity and history of previous caesarean section. Fourth, we examine “very small” size at birth as a proxy for preterm or growth-restricted newborns and evaluate whether any apparent survival differences could be explained by selective concentration of small or compromised fetuses into SBA or caesarean pathways. Finally, we translate the resulting absolute risk differences into simple scenario models to estimate the number of early neonatal deaths that might be averted each year if coverage of competent SBA and timely caesarean delivery were extended to poor and rural women who currently lack effective intrapartum care.

By combining nationally representative data on Nigerian facility births with rigorous, equity-focused causal methods, this work aims to clarify both how intrapartum care is distributed and what it buys in terms of early neonatal survival. In doing so, it seeks to inform health-system investments that move beyond increasing contact coverage toward reliably delivering the skilled intrapartum expertise and emergency obstetric capacity required to close the neonatal survival gap in high-burden settings.

### Research in context Evidence before this study

We searched PubMed, Embase, and the Demographic and Health Surveys (DHS) Program website up to March 31, 2025, using combinations of the terms *Nigeria*, *neonatal mortality*, *early neonatal death*, *skilled birth attendance*, *caesarean section*, *facility delivery*, and *propensity score* without language restrictions. We prioritised studies using nationally representative surveys or facility-based cohorts from low- and middle-income countries. Global estimates show that neonatal deaths now account for almost half of under-five mortality, with the highest burden in sub-Saharan Africa, including Nigeria. Existing DHS-based analyses from Nigeria and neighbouring countries suggest that caesarean section is often associated with higher crude neonatal mortality than vaginal birth, but these studies rely mainly on logistic regression or unadjusted comparisons and acknowledge substantial confounding by indication and socio-economic gradients in access to surgery. Similarly, evaluations of skilled birth attendance and facility delivery show inconsistent survival benefits once background risk and health-system quality are considered. We found no prior studies that: (1) focused specifically on facility-based singleton births in Nigeria; (2) quantified absolute risk differences and “deaths averted per 1,000 births” for both skilled attendance and caesarean section; and (3) used modern causal methods such as propensity-score overlap weighting to define an interpretable target population.

### Added value of this study

Using the 2018 Nigeria DHS, we constructed a nationally representative cohort of facility-based singleton births and jointly examined skilled birth attendance, caesarean delivery, early neonatal death, and “very small” size at birth. We applied propensity-score overlap weighting, combined with the complex survey design, to compare women for whom either exposure was clinically plausible, thereby reducing confounding by indication and socio-economic position. This approach yields directly interpretable absolute effects expressed as percentage-point risk differences and deaths per 1,000 births. We also decomposed the system into joint care patterns (non-skilled vaginal, skilled vaginal, skilled caesarean), explored effect modification by obstetric risk, and modelled equity gradients and illustrative policy scenarios. Our findings show that, within the region of covariate overlap, caesarean section neither markedly increases nor decreases early neonatal mortality, whereas skilled attendance mainly signals concentration of higher-risk pregnancies rather than intrinsic harm or benefit. The analysis separates survival effects from shifts in fetal growth and prematurity by examining very small size at birth as a secondary outcome.

### Implications of all the available evidence

Taken together with prior work, our results suggest that, in Nigeria and likely in similar health-system contexts, raising coverage of facility births, skilled attendance, and caesarean sections is necessary but not sufficient to substantially reduce early neonatal deaths. The central problems are who reaches facilities with skilled staff and surgical capacity, how quickly high-risk cases are recognised and referred, and whether perioperative and immediate neonatal care are of adequate quality. Policies should therefore focus on equitable access to timely, indicated caesarean delivery and robust intrapartum and newborn care (including resuscitation and early postnatal monitoring), particularly for poor and rural families, rather than on crude volume targets alone. Methodologically, this study illustrates how survey-based overlap weighting and absolute risk metrics can be used to interrogate the real-world performance of obstetric systems and to generate policy-relevant estimates that can be replicated across countries using standard DHS data.

## 2. Methods

### 2.1 Study design and data source

We conducted a cross-sectional analysis of nationally representative birth-history data from the 2018 Nigeria Demographic and Health Survey (NDHS). The NDHS is part of the global DHS Program, which uses standardized questionnaires, field procedures, and data processing protocols to enable cross-country comparisons in reproductive, maternal, newborn, and child health.

The 2018 NDHS employed a stratified, two-stage cluster sampling design. In the first stage, enumeration areas were selected with probability proportional to size within strata defined by state and urban–rural residence. In the second stage, a fixed number of households was selected within each cluster. All women aged 15–49 years in sampled households were eligible for an interviewer-administered questionnaire that included a complete retrospective birth history and detailed questions on antenatal, intrapartum, and postnatal care for recent births.

This analysis uses the NDHS “births recode” file, which contains one record for each live birth reported in the maternal birth histories, with linked maternal and household characteristics.

We followed the STROBE (Strengthening the Reporting of Observational Studies in Epidemiology) recommendations for reporting observational studies.

### 2.2 Study population

From the 2018 NDHS births recode, we first selected all live births occurring in the 5 years preceding the survey interview date. To align with the structure of the NDHS questionnaires and reduce recall bias, we then restricted the analysis to the most recent birth per woman within this 5-year window.

Because our research question focuses on health-system performance during facility-based childbirth, we further restricted the sample to births reported as occurring in a health facility (public or private). Multiple gestation births were excluded because of their substantially different risk profile and clinical management compared with singleton births.

To create a consistent analytic cohort for causal estimation, we excluded births with missing information on the primary exposures (skilled birth attendance and caesarean delivery), the primary outcome (early neonatal death), and any of the prespecified covariates required for propensity score estimation. This yielded a complete-case cohort of singleton, facility-based, recent births with full information on exposures, outcomes, and confounders. A flow diagram (Figure 1) documents all inclusion and exclusion steps.

**Figure 1.**
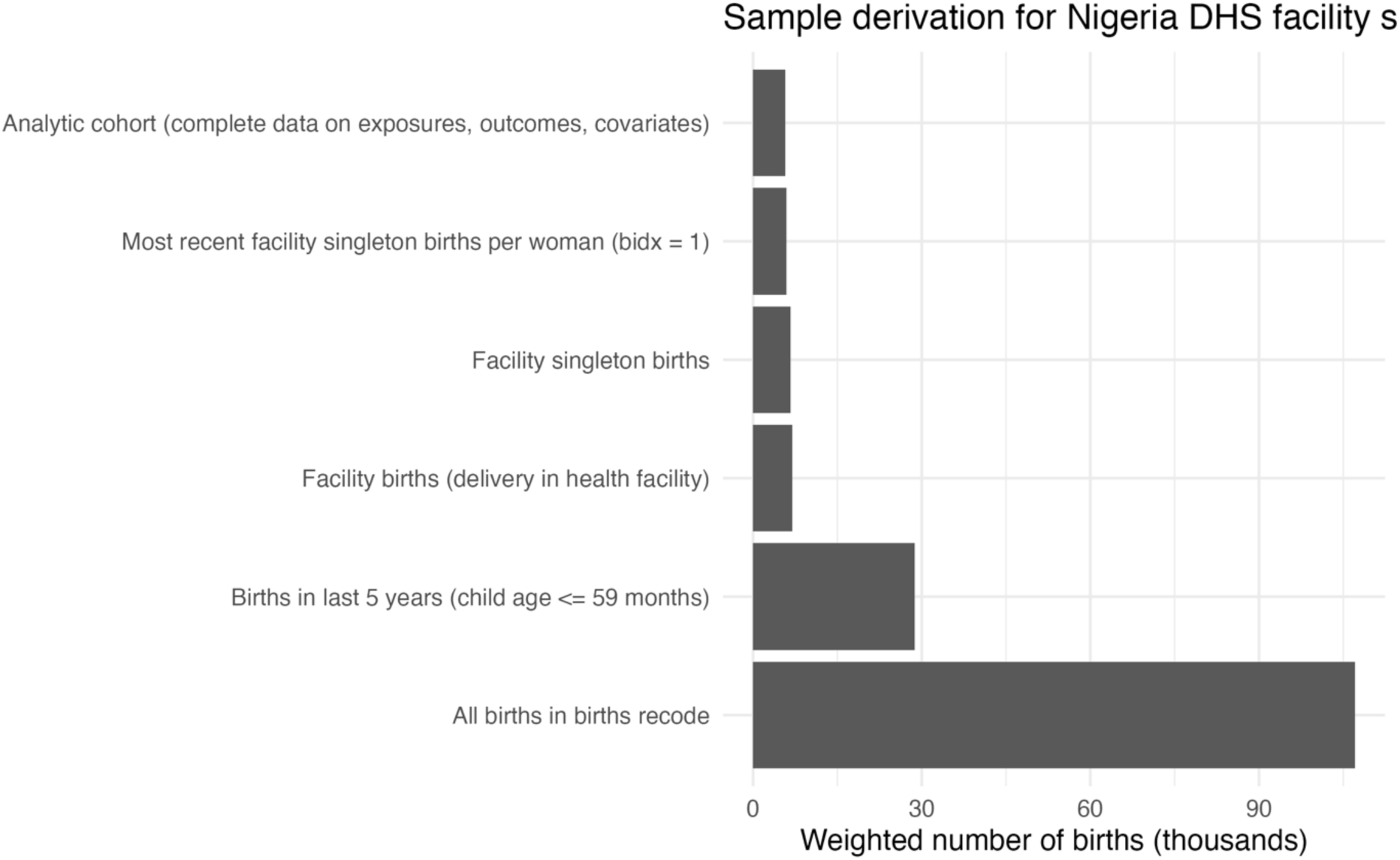
Sample derivation for the analytic cohort of facility-based singleton births, Nigeria DHS 2018. Bars show the survey-weighted number of births (in thousands) at each restriction step, from all births recorded in the births recode to births in the last 5 years, facility births, facility singleton births, most recent facility singleton birth per woman, and finally the analytic cohort of facility singleton births with complete data on exposures, outcomes, and covariates.

### 2.3 Exposures

We examined two main obstetric care exposures:

1. Skilled birth attendance (SBA). Skilled attendance at birth was defined using the standard DHS delivery-assistance variables, which capture up to three types of providers present at delivery. We coded skilled birth attendance as present if the woman reported that a doctor, nurse/midwife, or auxiliary midwife/health officer assisted during delivery, alone or in combination, and absent otherwise (including traditional birth attendants, relatives, and no one). This definition aligns with WHO and DHS classifications of “skilled health personnel” at birth.
2. Caesarean delivery. Mode of delivery was ascertained from the NDHS question on whether the baby was delivered by caesarean section. We coded caesarean delivery as a binary variable (yes vs. no).

For all analyses, facility type (public vs. private/other) was treated as a potential confounder rather than a primary exposure, because it strongly influences both the likelihood of receiving skilled care or caesarean section and neonatal outcomes in Nigeria.

### 2.4 Outcomes

#### Primary outcome: early neonatal death

The primary outcome was early neonatal death, defined among liveborn infants as death occurring between 0 and 6 completed days of life, consistent with WHO and DHS definitions of early neonatal mortality.

In the births recode file, we identified live births using the standard DHS survival status variable and calculated age at death in days. Early neonatal death was coded as 1 for infants who died between 0 and 6 days after birth and 0 for infants who survived at least 7 days (including those who later died in the late neonatal or post-neonatal period). Infants with implausible or missing information on age at death were excluded from the analytic cohort.

#### Secondary outcome: very small size at birth

As a secondary outcome, we examined maternal report of “very small size at birth,” a widely used DHS variable that serves as a proxy for severe fetal growth restriction or very low birthweight in settings where birthweight is often unmeasured.

Immediately after the questions on birthweight, women are asked to rate the baby’s size at birth on a 5-point scale (very large, larger than average, average, smaller than average, very small). We defined “very small size at birth” as a binary variable indicating that the mother reported the baby as “very small,” and 0 otherwise. Because this measure is subjective and may be influenced by maternal education and other social factors, we treated it as a secondary outcome and adjusted for an extensive set of socio-economic covariates in all analyses.

### 2.5 Covariates and causal framework

We selected covariates a priori using a directed acyclic graph (DAG) informed by prior literature on determinants of early neonatal mortality and access to obstetric care in low- and middle-income countries, as well as standard causal inference texts. The minimal adjustment set aimed to block back-door paths between obstetric care and neonatal outcomes without conditioning on mediators.

Covariates included:

- Maternal age at birth (years, modeled continuously or in categories for descriptive tables).
- Parity and birth order (number of previous live births; birth order of the index child).
- Maternal education (none, primary, secondary, higher).
- Household wealth quintile (DHS asset-based index, coded in quintiles).<u>PMC</u>
- Place of residence (urban vs. rural).
- Geopolitical region (six standard Nigerian regions or survey regions).
- Number of antenatal care (ANC) visits for the index pregnancy (0, 1–3, 4–7, ≥8).
- Infant sex (male vs. female).
- Birth year within the 5-year window before the survey (to capture secular trends).
- Facility sector of delivery (public vs. private/other).

These variables were included both in descriptive analyses and in propensity score models, reflecting their dual role as potential confounders of the relationship between facility-based care and neonatal outcomes and as key determinants of health-system equity in Nigeria.

### 2.6 Handling of survey design and weights

All analyses accounted for the complex survey design of the NDHS, including sampling weights, clustering, and stratification. We used the DHS individual sampling weight for births (v005), rescaled by dividing by 1,000,000 as per DHS guidance. Primary sampling units were defined by DHS clusters, and strata were defined using survey-specific stratification variables that combine state and urban–rural residence.

We constructed survey design objects using standard methods for complex survey analysis and applied these design specifications to all descriptive and regression analyses.

### 2.7 Descriptive analyses

We first described the distribution of maternal sociodemographic characteristics, obstetric care indicators, and neonatal outcomes in the complete-case analytic cohort, using survey-weighted means, proportions, and 95% confidence intervals (CIs). We compared characteristics of births included in the analytic cohort with those excluded due to missing data, to assess potential selection bias introduced by complete-case analysis.

To assess equity in coverage, we estimated survey-weighted prevalence of skilled birth attendance and caesarean delivery overall and stratified by wealth quintile, region, and urban–rural residence. For each proportion, we computed design-based 95% CIs using standard Taylor linearization methods.

### 2.8 Propensity score overlap weighting

To approximate a randomized comparison between exposure groups while retaining generalizability, we used propensity score overlap weighting (OW) for each primary exposure (skilled birth attendance and caesarean delivery). Overlap weights emphasize individuals with the highest probability of receiving either treatment (i.e., those in the region of covariate overlap) and have been shown to produce exact balance in means of observed covariates between exposure groups in large samples, with favorable small-sample properties.

For each exposure, we fit a logistic regression model for the probability of receiving the exposure (e.g., skilled attendance vs. no skilled attendance), conditional on all covariates listed above, using the original DHS sampling weights. Predicted probabilities were then used to compute overlap weights, such that individuals with extreme propensity scores received lower weights, and those in the overlapping region of the covariate distribution received higher weights. We multiplied these overlap weights by the original DHS sampling weights to construct final analysis weights and created new survey design objects based on the reweighted pseudo-population.

We assessed covariate balance before and after overlap weighting using absolute standardized mean differences (SMDs). An absolute SMD <0.10 was considered evidence of adequate balance between exposure groups.

### 2.9 Association between obstetric care and early neonatal death

We estimated the association between each obstetric care exposure and early neonatal death in two ways:

1. Survey-weighted models: We fit survey-weighted logistic regression models with early neonatal death as the dependent variable and each exposure as the main independent variable, adjusting for all covariates. These models provided adjusted odds ratios (aORs) and 95% CIs, accounting for clustering, stratification, and sampling weights.
2. Overlap-weighted models (primary causal contrast): Using the overlap-weighted survey design, we fit logistic regression models for early neonatal death as a function of the exposure alone, treating the OW procedure as having balanced confounders between exposure groups. These models approximated the average treatment effect in the overlap population.

For both sets of models, we obtained predicted probabilities of early neonatal death under observed exposure status and, using marginal standardization, under counterfactual scenarios in which all infants were exposed vs. all were unexposed. From these predicted risks, we calculated:

- Absolute risk differences (RDs) in deaths per birth;
- Relative risks (RRs); and
- Deaths averted (or excess deaths) per 1,000 births, computed as RD × 1,000.

Standard errors and 95% CIs for these derived measures were obtained using the delta method within the survey framework.

### 2.10 Secondary analyses: very small size at birth

We repeated the analytic strategy described above for the secondary outcome of very small size at birth. Specifically, we estimated survey-weighted and overlap-weighted associations between each exposure (skilled attendance, caesarean delivery) and the risk of very small size at birth, adjusting for the same set of covariates. Given the potential for reporting bias in maternal perception of birth size, we interpreted these results as exploratory and supportive of the main mortality findings rather than as definitive estimates of causal effect.

#### Sensitivity analyses

To examine the robustness of our findings, we conducted several prespecified sensitivity analyses:

1. Alternative covariate specifications. We re-estimated propensity scores using alternative coding of maternal age (categorical instead of continuous) and ANC visits (continuous instead of categorical), to assess the impact of functional form assumptions on overlap weighting and effect estimates.<u>eBay</u>
2. Restriction to higher-risk groups. We repeated analyses among subgroups defined by parity and maternal age (e.g., primiparous women, women aged ≥35 years) to explore whether associations differed in groups at higher baseline risk of adverse neonatal outcomes.
3. Complete survey-weighted models without overlap weighting. To demonstrate that our conclusions did not depend on the choice of weighting strategy, we compared overlap-weighted estimates with standard survey-weighted regression estimates adjusted for the same set of covariates.<u>PubMed+1</u>

All analyses were conducted using R (R Foundation for Statistical Computing) with packages for data management and complex survey analysis. Survey-weighted estimates accounted for clustering, stratification, and sampling weights in all stages of analysis.

### 2.11 Scenario modelling

To translate adjusted absolute effects into policy-relevant metrics, we combined overlap-weighted RDs with external estimates of the annual number of facility-based singleton births in Nigeria to approximate the number of early neonatal deaths potentially avertable under improved intrapartum care scenarios. In the main scenario, we assumed: (1) SBA coverage among facility births in the poorest wealth quintile and high-burden states increased to at least the national median; and (2) caesarean delivery was reliably available for all women with clearly indicated overlap-risk profiles. Averted deaths were calculated as the product of the RD in each stratum and the corresponding number of births, summing across strata. These estimates are intended as illustrative and depend on the assumptions that the overlap-weighted effects are causal and that scaling up care would deliver a similar quality of intrapartum and perioperative services.

### 2.12 Software

All analyses were conducted using RStudio version 2025.09.1+401, with dedicated survey and causal inference packages for PS estimation, overlap weighting, and variance estimation. Code is available from the corresponding author on reasonable request.

### 2.13 Ethical considerations

The 2018 NDHS protocol was approved by the National Health Research Ethics Committee of Nigeria and the ICF Institutional Review Board. Informed consent was obtained from all participating women before the interview (NPC & ICF, 2019). The anonymised, publicly available NDHS data were accessed with permission from The DHS Program. This secondary analysis of de-identified data was deemed exempt from additional institutional review board oversight according to prevailing guidance for analyses of existing public-use datasets.

## 3. RESULTS

### 3.1 Study population and outcome frequencies

In the 2018 Nigeria DHS births recode, 104,557 live births were reported by interviewed women. Restricting to births in the five years preceding the survey yielded 27,783 births. Of these, 7,245 occurred in a health facility, 6,870 were facility singleton births, and 6,072 were the most recent facility singleton birth per woman. After excluding observations with missing data on early neonatal survival, size at birth, skilled attendance, mode of delivery, or prespecified covariates, the final analytic cohort comprised 5,860 facility-based singleton births (Figure 1; Table 1). All sample counts are unweighted; all percentages and effect estimates are survey-weighted.

**Table 1.**
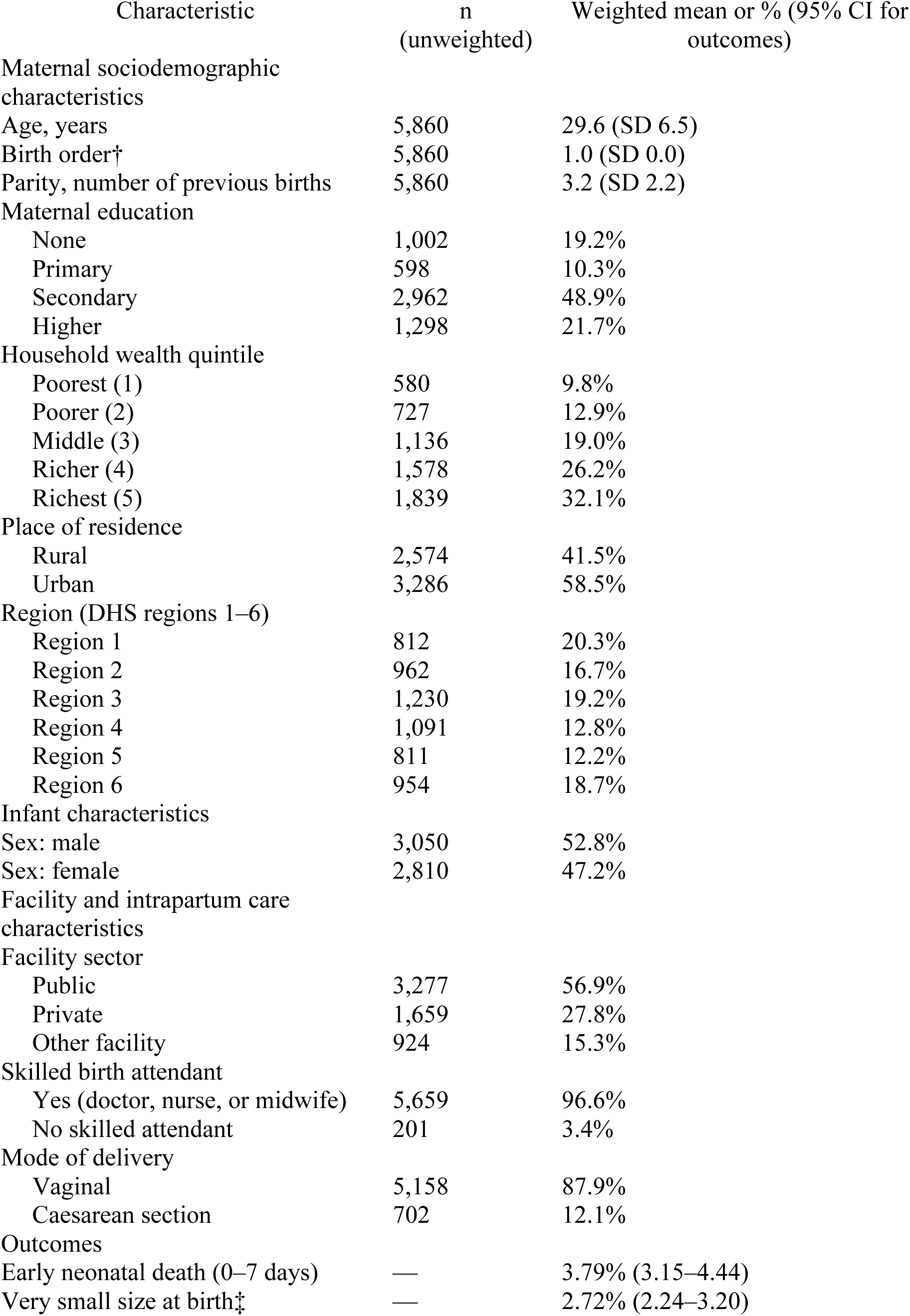
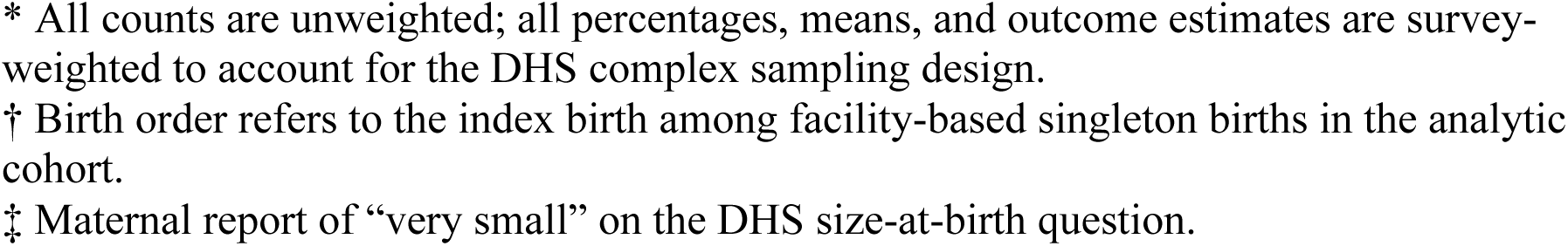
Baseline characteristics and outcomes among facility-based singleton births, Nigeria DHS 2018 (analytic cohort, n = 5,860*)

| Characteristic | n<br>(unweighted) | Weighted mean or % (95% CI for<br>outcomes) |
| --- | --- | --- |
| Maternal sociodemographic characteristics |  |  |
| Age, years | 5,860 | 29.6 (SD 6.5) |
| Birth order† | 5,860 | 1.0 (SD 0.0) |
| Parity, number of previous births | 5,860 | 3.2 (SD 2.2) |
| Maternal education |  |  |
| None | 1,002 | 19.2% |
| Primary | 598 | 10.3% |
| Secondary | 2,962 | 48.9% |
| Higher | 1,298 | 21.7% |
| Household wealth quintile |  |  |
| Poorest (1) | 580 | 9.8% |
| Poorer (2) | 727 | 12.9% |
| Middle (3) | 1,136 | 19.0% |
| Richer (4) | 1,578 | 26.2% |
| Richest (5) | 1,839 | 32.1% |
| Place of residence |  |  |
| Rural | 2,574 | 41.5% |
| Urban | 3,286 | 58.5% |
| Region (DHS regions 1–6) |  |  |
| Region 1 | 812 | 20.3% |
| Region 2 | 962 | 16.7% |
| Region 3 | 1,230 | 19.2% |
| Region 4 | 1,091 | 12.8% |
| Region 5 | 811 | 12.2% |
| Region 6 | 954 | 18.7% |
| Infant characteristics |  |  |
| Sex: male | 3,050 | 52.8% |
| Sex: female | 2,810 | 47.2% |
| Facility and intrapartum care characteristics |  |  |
| Facility sector |  |  |
| Public | 3,277 | 56.9% |
| Private | 1,659 | 27.8% |
| Other facility | 924 | 15.3% |
| Skilled birth attendant |  |  |
| Yes (doctor, nurse, or midwife) | 5,659 | 96.6% |
| No skilled attendant | 201 | 3.4% |
| Mode of delivery |  |  |
| Vaginal | 5,158 | 87.9% |
| Caesarean section | 702 | 12.1% |
| Outcomes |  |  |
| Early neonatal death (0–7 days) | — | 3.79% (3.15–4.44) |
| Very small size at birth‡ | — | 2.72% (2.24–3.20) |

\* All counts are unweighted; all percentages, means, and outcome estimates are survey-weighted to account for the DHS complex sampling design.
† Birth order refers to the index birth among facility-based singleton births in the analytic cohort.
‡ Maternal report of “very small” on the DHS size-at-birth question.

After application of DHS sampling weights, the analytic cohort remained nationally representative of Nigerian facility births. The weighted prevalence of early neonatal death (0–7 days) was 3.8% (95% CI 3.2–4.4), and 2.7% (95% CI 2.2–3.2) of neonates were reported as “very small” at birth (Table 1; Supplementary Figure S1).

Mothers had a mean age of 29.6 years (SD 6.5) and a median of 29 years (IQR 25–34). Median parity was 3 (IQR 2–5). Educational attainment was heterogeneous: 19% had no schooling, 10% had primary, 49% had secondary, and 22% had higher education. Births were distributed across wealth quintiles (approximately 10%, 13%, 19%, 26%, and 32% from the poorest to richest quintile), and 41.5% occurred in rural areas.

Most deliveries took place in public facilities (56.9%), followed by private facilities (27.8%) and other facility types (15.3%). The sex distribution of newborns was balanced (52.8% male, 47.2% female). Skilled birth attendance (SBA) was nearly universal: 96.6% of births were attended by a doctor, nurse, or midwife, and 3.4% lacked a skilled attendant despite occurring in a facility. Caesarean section accounted for 12.1% of deliveries, with the remainder vaginal births. All subsequent estimates are based on this analytic cohort and incorporate survey design features and sampling weights to yield nationally interpretable estimates for facility-based singleton births.

### 3.2 Distribution of obstetric risk, skilled care, and caesarean section

Obstetric and socio-economic risk were strongly patterned across skilled attendance and caesarean delivery among facility singleton births. Despite high overall SBA coverage (>94%), the small group of births without a skilled attendant (201/5,860) were markedly disadvantaged. Approximately 60% of non-skilled births occurred in rural areas (vs ∼41% of skilled births), and 43% fell in the two poorest wealth quintiles (vs 22% among skilled births). In contrast, births with SBA were concentrated in the top two wealth quintiles (59% vs 29% of non-skilled births) and in public or private hospitals rather than lower-level “other” facilities (86% vs 52%). Thus, non-skilled attendance within facilities was rare but concentrated at the bottom of the socio-economic gradient.

Caesarean delivery showed an even sharper socio-economic gradient. Overall, 12.1% of facility singleton births were delivered by caesarean section (702/5,860). Compared with vaginal births, women undergoing C-section were older (30.9 vs 29.4 years), more often urban (71% vs 57%), and substantially more likely to be in the richest wealth quintile (52% vs 29%). Only about 4% of C-section births occurred in the poorest quintile, compared with 11% of vaginal births. Caesarean deliveries were also shifted towards private facilities: 46% of C-sections versus 25% of vaginal births occurred in private facilities, whereas only 3% versus 17%, respectively, occurred in lower-level “other” facilities (Table 2).

Obstetric risk factors were similarly skewed. Among women with a prior caesarean, roughly three-quarters delivered again by C-section. Higher education, greater antenatal care (ANC) use, and urban residence clustered among women receiving SBA or C-section, whereas women without SBA or with vaginal delivery were more likely to have lower schooling and higher parity.

Before adjustment, these gradients generated substantial confounding by indication. Absolute standardised mean differences between exposure groups frequently exceeded 0.3–0.4 for wealth, facility sector, region, and urban residence, and were substantial for maternal age, parity, and ANC use. After applying propensity-score overlap weights, all prespecified covariates for both the SBA and C-section models had absolute standardised differences close to zero and below 0.1, indicating excellent balance and supporting use of the overlap-weighted contrasts in subsequent analyses (Figure 2B).

**Figure 2B.**
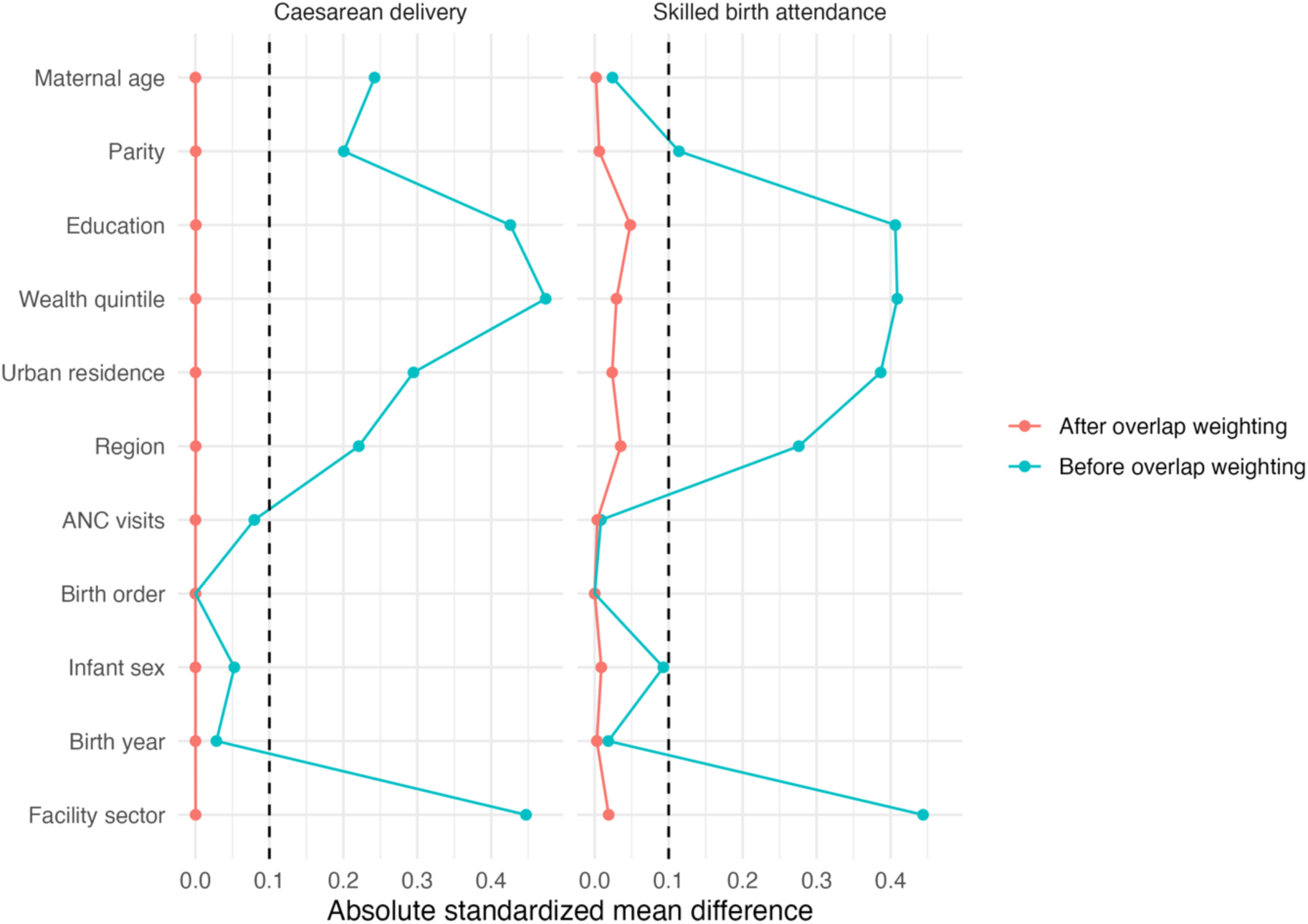
Covariate balance before and after propensity-score overlap weighting for caesarean delivery and skilled birth attendance. Absolute standardized mean differences are shown for each covariate before (blue) and after (red) overlap weighting in the caesarean and SBA models. The dashed vertical line at 0.10 marks the conventional threshold for acceptable imbalance. Before weighting, several covariates—particularly wealth quintile, region, urban residence, education, and facility sector—had substantial imbalance (absolute standardized mean differences often >0.30–0.40). After overlap weighting, all covariates in both models were reduced to <0.10, indicating excellent balance and supporting the validity of the overlap-weighted contrasts.

### 3.3 Skilled birth attendance and early neonatal death

Comparisons between births with and without SBA were strongly confounded by indication, with births attended by skilled providers drawn from systematically higher-risk women.

In crude survey-weighted analyses parameterised as “SBA vs non-SBA,” births with SBA had a higher risk of early neonatal death (3.9%) than those without SBA (1.7%), corresponding to an absolute risk difference of 2.2 percentage points (95% CI −0.1 to 4.4) and a risk ratio of 2.26 (95% CI 0.63–8.16) (Table 3). This pattern is consistent with strong confounding by indication and facility case-mix rather than harm from skilled attendance.

**Table 3.**
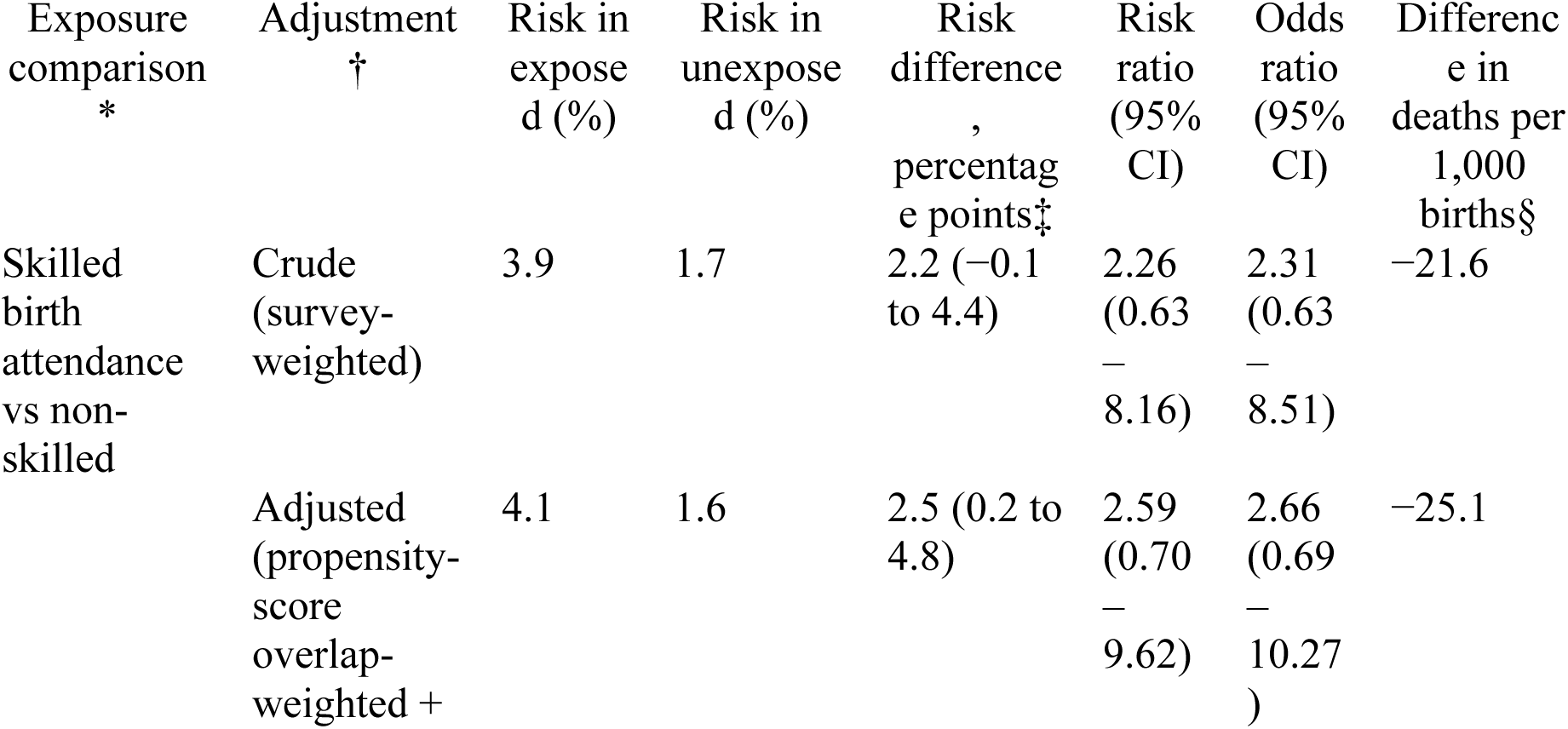
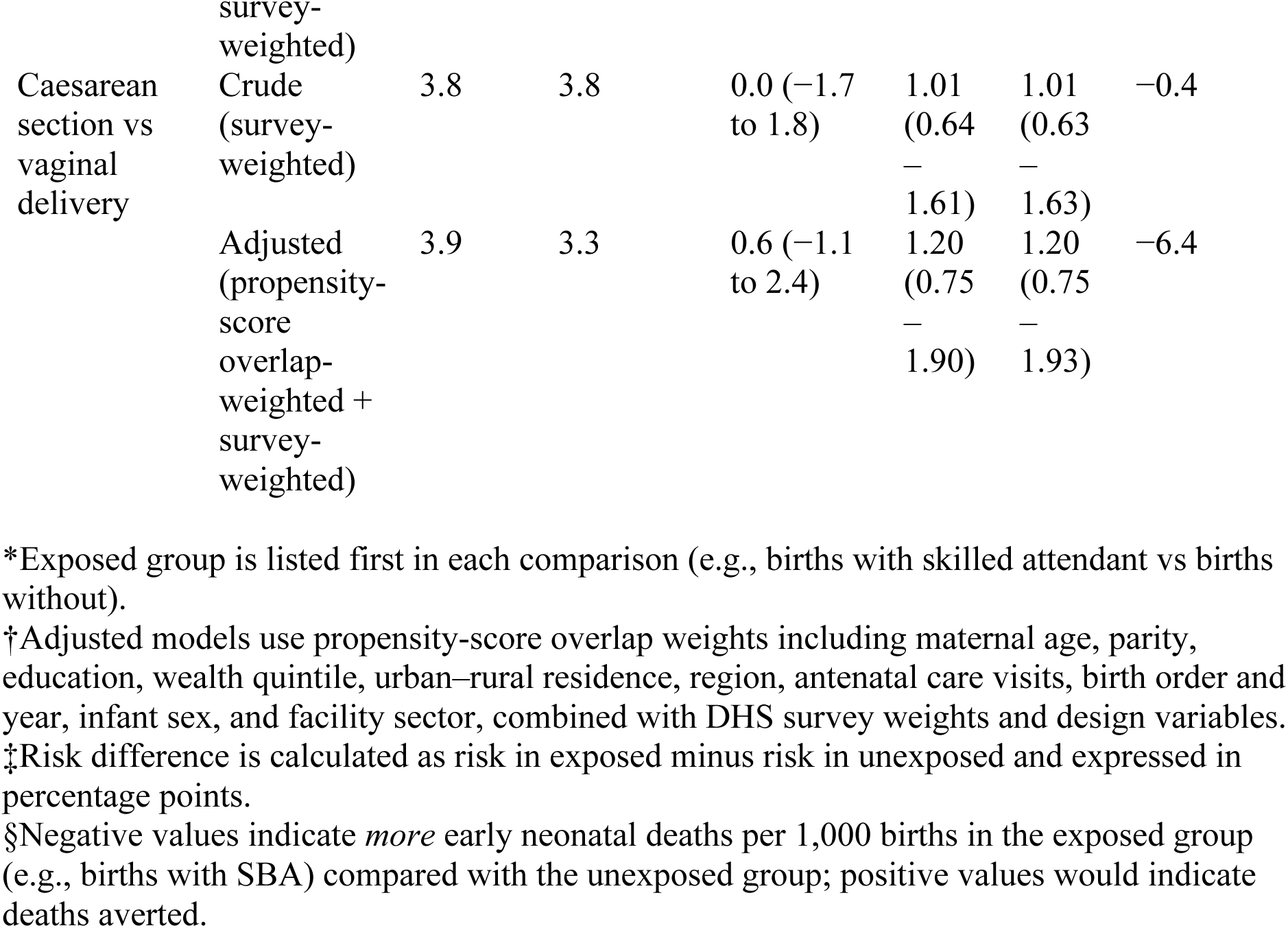
Crude and adjusted associations of skilled birth attendance and caesarean delivery with early neonatal death among facility-based singleton births, Nigeria DHS 2018

After applying overlap weighting on the DHS survey design to balance maternal age, parity, education, wealth quintile, urban–rural residence, region, ANC visits, birth order and year, infant sex, and facility sector, the direction of the association remained similar and the contrast slightly increased. In the overlap population, the risk of early neonatal death was 4.1% among births with SBA and 1.6% among those without, yielding an adjusted risk difference of 2.5 percentage points (95% CI 0.2–4.8). On the absolute scale, this corresponds to about 25 additional deaths per 1,000 facility singleton births when comparing the observed SBA group with the small group who did not receive SBA. Re-expressed as “no SBA versus SBA,” these estimates imply that failing to secure a skilled attendant is associated with approximately 22–25 more early neonatal deaths per 1,000 births among otherwise comparable women, although precision is limited.

On the relative scale, the overlap-weighted adjusted risk ratio for early neonatal death comparing SBA with non-SBA was 2.59 (95% CI 0.70–9.62), and the corresponding odds ratio was 2.66 (95% CI 0.69–10.27). Confidence intervals were wide and compatible with sizeable harm or modest benefit on the relative scale, reflecting the rarity of non-skilled facility births and the limited number of deaths in this group. Across crude and adjusted models, point estimates consistently suggested that, under the observed allocation of care, births attended by skilled providers were drawn from higher-risk women than births without SBA—a pattern more plausibly attributable to residual confounding by unmeasured obstetric complexity, intrapartum complications, and referral pathways than to a true causal hazard from skilled attendance.

Crude and overlap-weighted risk differences are shown in Figure 3A; the full set of risks, risk differences, risk ratios, and odds ratios is presented in Table 3 and Supplementary Table S1.

**Figure 3A.**
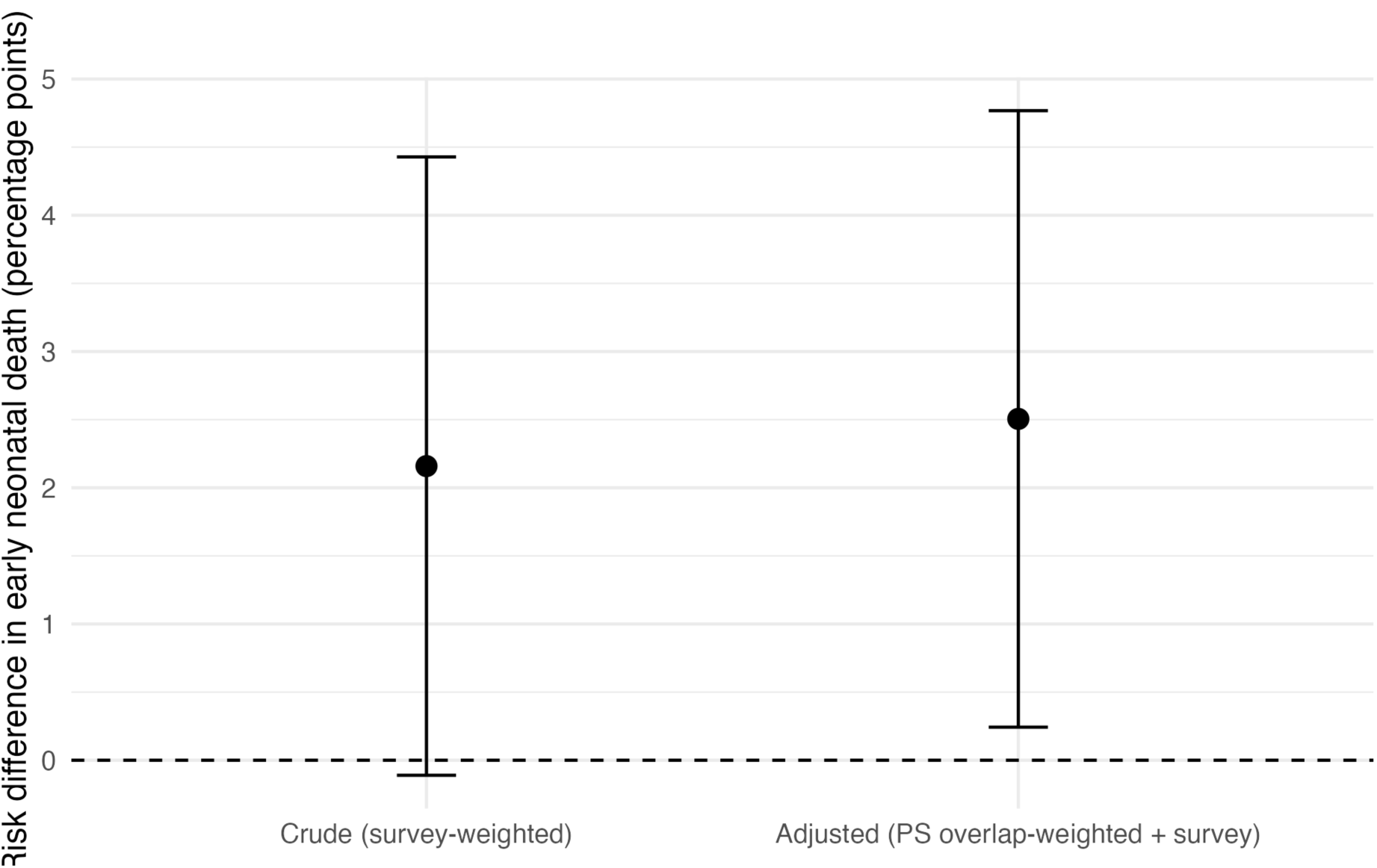
Crude and overlap-weighted risk differences in early neonatal death for skilled vs non-skilled birth attendance among facility-based singleton births, Nigeria DHS 2018. Points show absolute risk differences (percentage points) comparing births with skilled attendance to births without skilled attendance; error bars indicate 95% confidence intervals from survey-weighted (crude) and propensity-score overlap-weighted models. Positive values indicate higher early neonatal mortality among births with skilled attendance relative to non-skilled attendance.

### 3.4 Caesarean section and early neonatal death

Within clinically comparable women who could plausibly deliver either vaginally or by caesarean section, there was no compelling evidence that C-section substantially increases or decreases early neonatal mortality.

In crude survey-weighted analyses, the risk of early neonatal death was essentially identical for caesarean and vaginal births (3.8% vs 3.8%; risk difference 0.0 percentage points, 95% CI −1.7 to 1.8; risk ratio 1.01, 95% CI 0.64–1.61; Table 4). These unadjusted estimates provide little evidence of large overall harm or benefit from C-section when confounding by indication is not addressed.

**Table 4.**
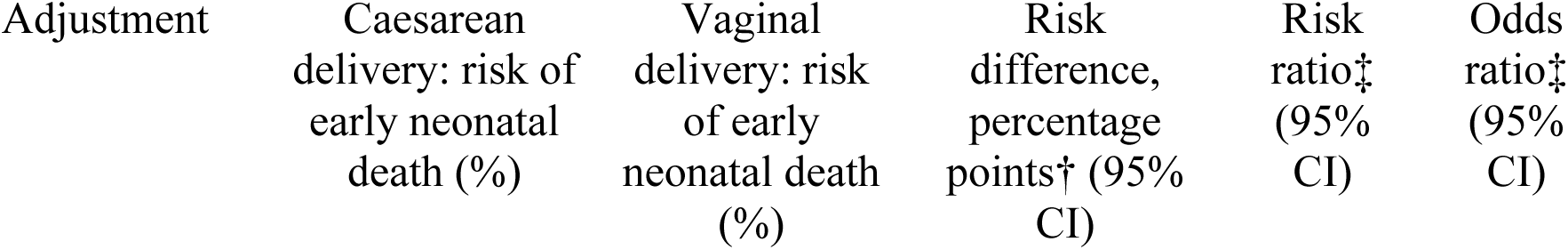
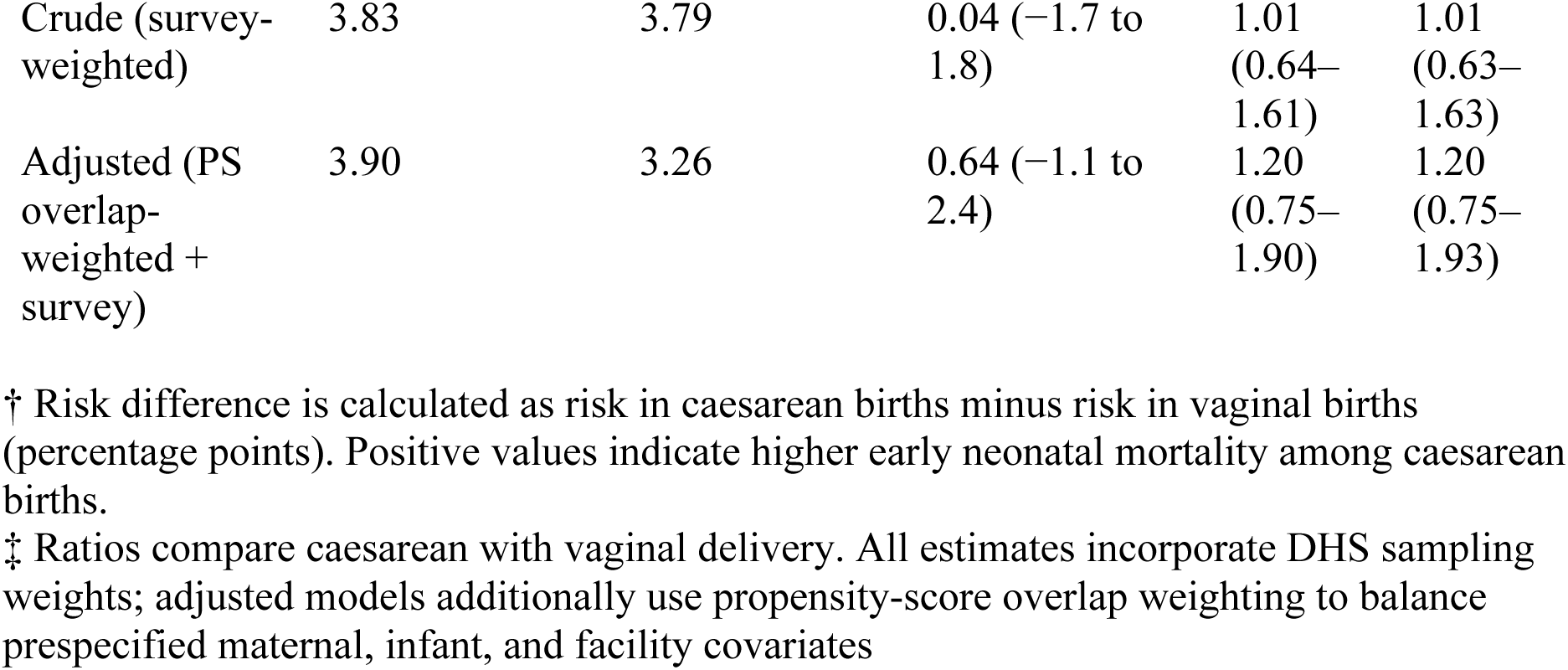
Crude and adjusted associations of caesarean delivery with early neonatal death among facility-based singleton births, Nigeria DHS 2018.

| Adjustment | Caesarean delivery: risk of early neonatal death (%) | Vaginal delivery: risk of early neonatal death (%) | Risk difference, percentage points† (95% CI) | Risk ratio‡ (95% CI) | Odds ratio‡ (95% CI) |
| --- | --- | --- | --- | --- | --- |
| Crude (survey-weighted) | 3.83 | 3.79 | 0.04 (−1.7 to 1.8) | 1.01 (0.64–1.61) | 1.01 (0.63–1.63) |
| Adjusted (PS overlap-weighted + survey) | 3.90 | 3.26 | 0.64 (−1.1 to 2.4) | 1.20 (0.75–1.90) | 1.20 (0.75–1.93) |

† Risk difference is calculated as risk in caesarean births minus risk in vaginal births (percentage points). Positive values indicate higher early neonatal mortality among caesarean births.
‡ Ratios compare caesarean with vaginal delivery. All estimates incorporate DHS sampling weights; adjusted models additionally use propensity-score overlap weighting to balance prespecified maternal, infant, and facility covariates

After overlap weighting on maternal age, parity, education, wealth quintile, urban–rural residence, region, ANC use, birth order and year, infant sex, and facility sector, point estimates moved modestly away from the null. In the overlap population—women whose clinical and socio-demographic characteristics make either mode of delivery plausible—the risk of early neonatal death was 3.9% among caesarean births and 3.3% among vaginal births (risk difference 0.6 percentage points, 95% CI −1.1 to 2.4; risk ratio 1.20, 95% CI 0.75–1.90; adjusted odds ratio 1.20, 95% CI 0.75–1.93). The confidence intervals span both small protective and small harmful effects, indicating no statistically compelling evidence that C-section either increases or decreases early neonatal mortality within the region of covariate overlap.

Expressed on an absolute scale, the adjusted risk difference corresponds to approximately 6 additional early neonatal deaths per 1,000 caesarean births compared with vaginal births, with uncertainty ranging from about 11 fewer to 24 additional deaths per 1,000 births (Figure 4B). These bounds rule out large detrimental effects of medically indicated C-section among Nigerian facility births, while suggesting that any survival advantage is likely to be modest and dependent on timely, high-quality perioperative and neonatal care.

Taken together with the strong socio-economic gradients in C-section access, these findings are consistent with crude excess mortality among C-section births reflecting concentration of the most complicated labours and fetuses into surgical pathways rather than intrinsic harm from surgery. Crude and overlap-weighted risk differences are shown in Figure 4A; the full set of estimates is given in Table 4 and Supplementary Table S1.

**Figure 4A.**
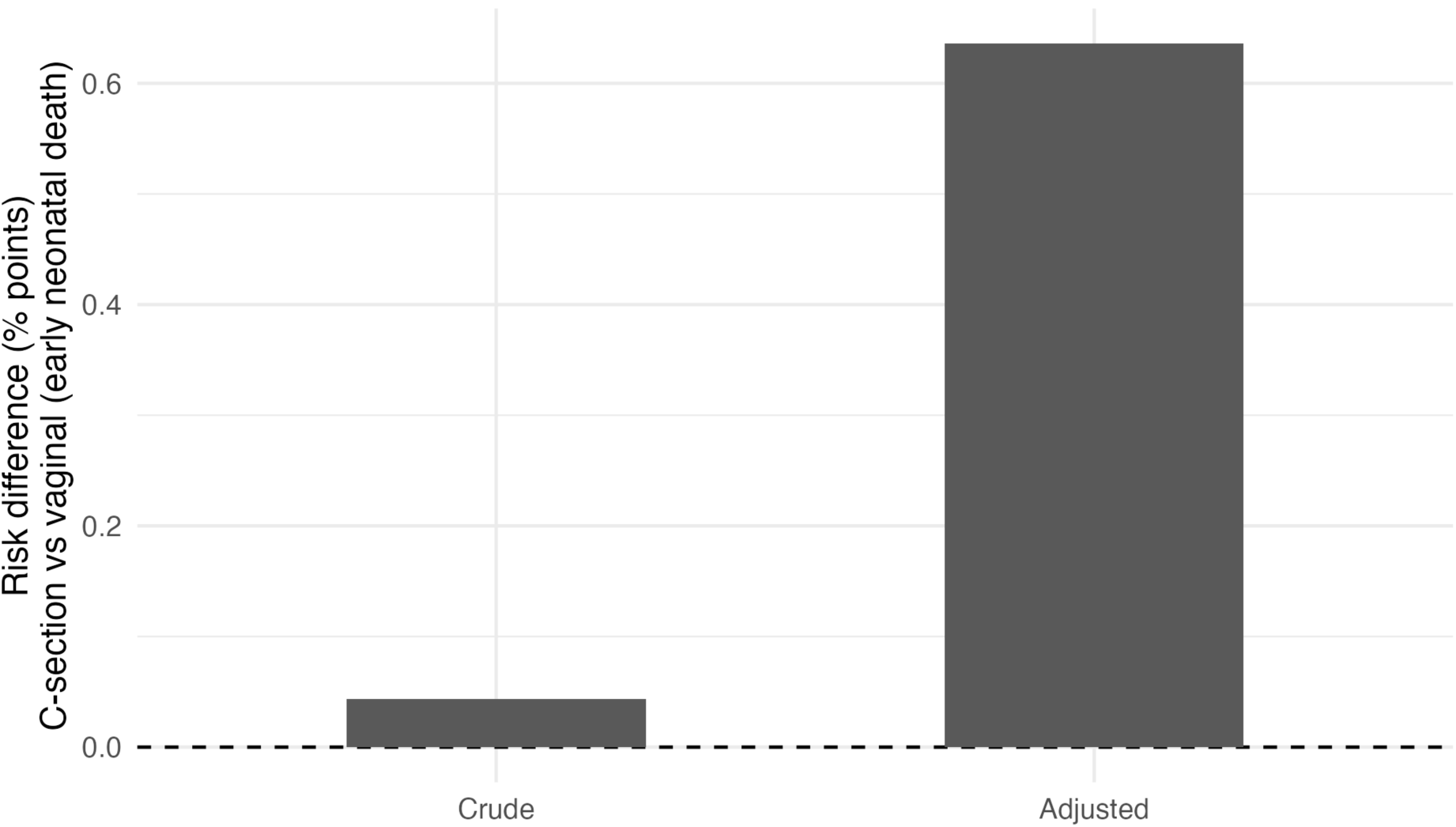
Crude and overlap-weighted risk differences for early neonatal death comparing caesarean section with vaginal delivery among facility-based singleton births, Nigeria DHS 2018.

Bars show the absolute risk difference in early neonatal death (percentage-point difference in risk, caesarean minus vaginal delivery), estimated using crude survey-weighted models and propensity-score overlap-weighted models that also incorporate the DHS survey design. Positive values indicate higher early neonatal death risk following caesarean delivery compared with vaginal birth.

### 3.5 Very small size at birth

Skilled intrapartum care was not strongly associated with shifts in the distribution of very small size at birth, suggesting that survival patterns are not explained solely by selective allocation of preterm or growth-restricted fetuses into SBA or C-section pathways.

For SBA, crude survey-weighted analyses suggested a slightly higher prevalence of very small size among births with SBA (2.8%) than among those without SBA (1.6%), corresponding to a crude risk difference of 1.2 percentage points (95% CI −1.1 to 3.5) and a risk ratio of 1.75 (95% CI 0.42–7.23; Table 5). After overlap weighting on maternal age, parity, education, wealth, urban residence, region, ANC visits, birth order and year, infant sex, and facility sector, the difference narrowed: very small size remained uncommon in both groups (2.4% with SBA vs 1.6% without SBA), yielding an adjusted risk difference of 0.7 percentage points (95% CI −1.7 to 3.2) and an odds ratio of 1.47 (95% CI 0.33–6.48).

Patterns were similar for caesarean delivery. In crude analyses, very small size was slightly more frequent among C-sections (3.6%) than vaginal births (2.6%), with a risk difference of 1.0 percentage point (95% CI −0.6 to 2.7) and a risk ratio of 1.40 (95% CI 0.87–2.26). After overlap weighting, absolute risks remained low (3.5% vs 2.7%), and the adjusted risk difference was 0.9 percentage points (95% CI −0.8 to 2.5), with an odds ratio of 1.35 (95% CI 0.80–2.28).

Figure 5A summarises these crude and overlap-weighted risk differences for SBA and C-section side-by-side. In both contrasts, shifts in the distribution of very small size are small and statistically uncertain, especially when compared with the larger absolute differences observed for early neonatal death. Ordered-category analyses across all DHS size groups (very large to very small) likewise showed no strong monotonic shift toward smaller size after adjustment (Supplementary Figure S5; Supplementary Table S5). These findings are therefore more consistent with skilled intrapartum care acting primarily through intrapartum and immediate neonatal management than through systematic reallocation of growth-restricted or preterm fetuses into specific delivery modes.

### 3.6 Effect modification, equity gradients, and system-level patterns

#### 3.6.1 Effect modification by obstetric risk

Associations between mode of delivery and early neonatal death varied only modestly across key obstetric risk strata and were estimated imprecisely. Among primiparous women (parity 0–1), overlap-weighted risk differences comparing C-section with vaginal delivery were directionally protective, with fewer early neonatal deaths per 1,000 births than among multiparous women, although confidence intervals were wide and included the null in both strata (Supplementary Table S7). Among multiparous women (parity ≥2), overlap-weighted risk differences suggested little or no benefit of C-section, consistent with many procedures being performed for softer indications or later in the course of fetal compromise.

When stratified by history of prior C-section, repeat caesarean was associated with early neonatal risks that were similar to or slightly lower than for attempted vaginal birth, again with substantial statistical imprecision due to small numbers. These patterns align with clinical expectations that the clearest neonatal survival gains from C-section occur in high-risk, clearly indicated scenarios.

For SBA, overlap-weighted effect estimates were remarkably stable across parity and previous C-section strata (Supplementary Table S7), suggesting that the principal margin of benefit is the presence of any skilled attendant rather than strong interaction with specific obstetric risk profiles.

#### 3.6.2 Socio-economic and geographic gradients

Facility-based early neonatal mortality exhibited a steep equity gradient: risks were highest in the poorest wealth quintiles, among rural residents, and in births occurring in lower-level public facilities. These same groups also had the lowest coverage of both C-section and SBA (Supplementary Table S8).

After overlap weighting, adjusted risk differences for C-section versus vaginal delivery were broadly similar across wealth quintiles and urban–rural residence, indicating that once women reach comparable facilities, the intrinsic neonatal effect of C-section does not differ materially by socio-economic status. The central inequity therefore lies in access and timeliness—who reaches skilled providers and timely surgery—rather than in differential effectiveness of these interventions by wealth or residence.

#### 3.6.3 Joint care patterns and system behaviour

To summarise intrapartum care as a system, we defined a three-level joint care pattern combining attendance and mode of delivery:

1. Non-skilled vaginal birth
2. Skilled vaginal birth
3. Skilled caesarean section

Survey-weighted risks of early neonatal death differed meaningfully across these patterns (Figure 6; Supplementary Table S9). Skilled vaginal births had the lowest mortality. Non-skilled vaginal births and skilled C-section births both showed higher risks, with skilled C-section births reflecting the highest concentration of obstetric complexity.

**Figure 6.**
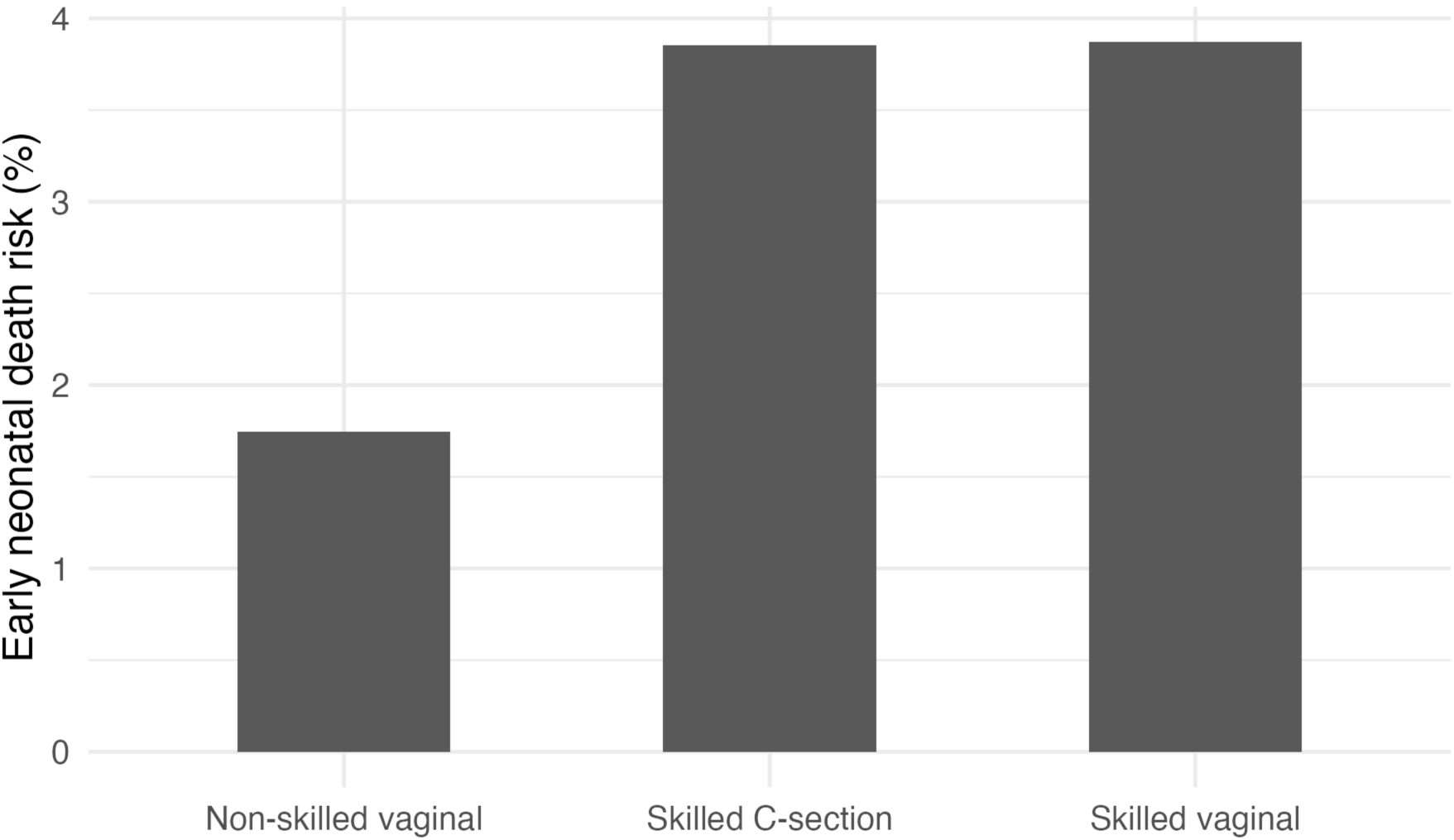
Gradient of early neonatal mortality by joint intrapartum care pattern, Nigeria DHS 2018. Survey-weighted risk of early neonatal death (0–7 days) among facility-based singleton births, shown for three joint care patterns: non-skilled vaginal birth, skilled vaginal birth, and skilled caesarean section. Bars show point estimates; sample is restricted to the most recent facility-based singleton birth per woman in the five years preceding the survey (unweighted n=5 860); all estimates incorporate DHS sampling weights and survey design.

**Table 6.** Facility-based early neonatal mortality and intrapartum care coverage by wealth quintile, Nigeria DHS 2018.

| Wealth quintile* | Unweighted births (n) | Weighted births (thousands) | Early neonatal deaths per 1,000 births | Risk difference vs wealthiest† (per 1,000) | Skilled birth attendance (%) | Caesarean delivery (%) |
| --- | --- | --- | --- | --- | --- | --- |
| 1 (Poorest) | 580 | 560 | 43.1 | +12.9 | 93.3 | 5.9 |
| 2 | 796 | 735 | 43.8 | +13.6 | 96.2 | 7.4 |
| 3 | 1,138 | 1,037 | 42.8 | +12.6 | 96.8 | 9.0 |
| 4 | 1,538 | 1,351 | 37.6 | +7.4 | 97.6 | 13.4 |
| 5<br>(Richest) | 1,808 | 1,584 | 30.2 | 0<br>(reference) | 98.8 | 21.9 |

\*Wealth quintiles are based on the DHS household asset index.
†Positive values indicate higher early neonatal mortality compared with the wealthiest quintile.

This configuration reflects two features of the current system. First, risk concentration: fetuses and mothers with the highest clinical risk are only partially and unevenly channelled into skilled caesarean delivery, particularly in urban and wealthier strata. Second, incomplete risk reduction: even with skilled personnel and surgery, survival among high-risk newborns remains substantially worse than among low-risk births attended by skilled providers, suggesting residual gaps in timeliness, perioperative care, neonatal resuscitation capacity, and postnatal monitoring.

#### 3.6.4 Negative-control and robustness analyses

As a negative-control outcome, we examined receipt of BCG vaccination at birth, an intervention influenced by postnatal care but not by intrapartum mode of delivery. After overlap weighting, C-section showed no meaningful association with BCG uptake, suggesting that the models are not merely capturing general differences in care-seeking or access to child health services.

Sensitivity analyses using alternative operational definitions of skilled attendance, trimming extreme propensity scores, and restricting the early neonatal window to 0–3 days yielded effect estimates similar in magnitude and direction to the primary analyses (Supplementary Tables S10–S12), supporting the robustness of the main conclusions.

#### 3.6.5 Scenario modelling and potentially avoidable deaths

Finally, we translated adjusted absolute effects into illustrative policy scenarios. Under a scenario in which (1) SBA coverage among facility births in the poorest wealth quintile and in high-burden states was raised to at least the current national median, and (2) caesarean delivery was reliably available for all women in clearly indicated overlap-risk profiles, modelled early neonatal mortality among facility singleton births declined meaningfully, with most of the avertable deaths concentrated among poor and rural families. The detailed projections are presented in the Supplementary Appendix and should be interpreted as approximations under stated assumptions rather than forecasts.

Figure 6 summarises the gradient in early neonatal mortality across joint care patterns; stratified and scenario results are shown in Supplementary Tables S7–S9 and Supplementary Figures S6B–S6C.

## 4. DISCUSSION

In this nationally representative cohort of 5,860 facility-based singleton births in Nigeria, early neonatal mortality remained high at 3.8% (95% CI 3.2–4.4) despite skilled birth attendance coverage of 96.6% and a caesarean rate of 12.1%. Using survey-weighted overlap propensity scores, we found no large survival advantage associated with skilled attendance or caesarean delivery as they are currently delivered. In the overlap population—women whose characteristics made either mode of delivery plausible—the risk of early neonatal death was 3.9% after caesarean section and 3.3% after vaginal birth, corresponding to an adjusted risk difference of 0.6 percentage points (95% CI −1.1 to 2.4), or about 6 additional deaths per 1,000 births, with confidence intervals compatible with modest harm or modest benefit. These findings suggest that in this setting, the promise of intrapartum care has not yet translated into reliable survival gains for newborns.

The unexpected pattern in which births with skilled attendance had higher mortality than those without—4.1% versus 1.6% after overlap weighting (risk difference 2.5 percentage points, 95% CI 0.2–4.8)—is best interpreted as a marker of selective referral and late presentation rather than harm from skilled care. Facility births without a skilled attendant were rare and concentrated among poorer, rural women in lower-level facilities; those with skilled providers were more likely to be older, urban, wealthier, and to have higher antenatal contact, characteristics consistent with a higher burden of clinical risk and with referral of complicated labours. In such systems, reaching a skilled provider is often a sign that complications have already evolved, compressing the window in which intrapartum care can avert death.

Analyses of very small size at birth reinforce this interpretation. Only 2.7% of neonates were reported as “very small”, and overlap-weighted differences in this proxy for extreme prematurity or severe growth restriction were small for both skilled attendance and caesarean delivery. Very small size remained uncommon across exposure groups (for caesarean vs vaginal, 3.5% vs 2.7%; adjusted risk difference 0.9 percentage points, 95% CI −0.8 to 2.5). The survival gradient is therefore unlikely to be driven solely by selective concentration of the smallest or sickest fetuses into particular care pathways. Instead, the data point towards intrapartum and immediate neonatal processes—timeliness of decision-making, quality of monitoring, access to safe surgery, neonatal resuscitation, and early postnatal care—as pivotal determinants of early neonatal outcomes.

Viewing intrapartum care as joint patterns rather than isolated interventions provides a clearer picture of system behaviour. When care is summarised as non-skilled vaginal birth, skilled vaginal birth, and skilled caesarean section, a consistent gradient emerges: non-skilled vaginal births have the highest early neonatal mortality; skilled vaginal births have the lowest; and skilled caesarean births fall in between. This configuration is coherent with substantial risk concentration into surgical pathways and incomplete risk reduction even after surgery. It suggests that caesarean section is averting some deaths among high-risk pregnancies, but that delays and weaknesses in perioperative and neonatal care prevent outcomes in this group from approaching those of low-risk births attended by skilled providers.

Equity patterns deepen this concern. Early neonatal mortality is highest among poor, rural families and births in lower-level public facilities—the same groups with the lowest coverage of caesarean section and, to a lesser extent, skilled attendance. Yet once women with similar characteristics reach comparable facilities, the overlap-weighted association between caesarean delivery and early neonatal death is broadly similar across wealth and urban–rural strata. This implies that the major inequity is not that caesarean section works differently for poor versus rich women, but that poorer women are less likely to reach timely, functional surgical and neonatal care. Policies that focus solely on raising national averages for caesarean or skilled attendance coverage, without addressing the distribution and quality of services, are unlikely to substantially reduce early neonatal deaths.

Stratified analyses hint at where gains are most likely. Among primiparous women and those with a history of caesarean, effect estimates for surgery were compatible with small survival benefits, although imprecision was substantial. Among multiparous women without clear high-risk indications, the association between caesarean section and early neonatal death was closer to the null, consistent with procedures performed late, for weaker indications, or in already compromised pregnancies. The negative-control analysis, in which caesarean section showed no association with BCG vaccination after overlap weighting, supports the view that our models are capturing a clinically plausible relationship with neonatal survival rather than simply reflecting general care-seeking behaviour.

This study has limitations inherent to observational analyses. Residual confounding by unmeasured obstetric complexity and facility-level readiness is likely, maternal reports of timing of death and size at birth are imperfect, and we cannot distinguish elective from emergency caesareans or fully characterise referral delays and decision-to-delivery intervals. The number of deaths in some strata is modest, leading to wide confidence intervals. Nonetheless, the use of a recent, nationally representative survey; restriction to singleton facility births; formal handling of the complex survey design; and application of overlap weighting to focus inference on clinically meaningful contrasts together provide a robust description of how current intrapartum care in Nigeria is performing where it matters most: in the first week of life.

Taken together, these findings suggest that Nigeria’s challenge is not the absence of efficacious intrapartum interventions, but a misalignment between risk, access, and readiness. Skilled personnel and surgical capacity are present, but too many high-risk women reach them too late, and too many facilities lack the capability to deliver integrated obstetric and neonatal care at the moment of need. Future work should treat intrapartum care as a system, quantifying how coordinated improvements in triage, referral, surgical availability, and neonatal care—especially in poor and rural settings—could shift the distribution of care patterns and drive early neonatal mortality well below current levels.

## 5. CONCLUSION

In a large, nationally representative cohort of Nigerian facility births, early neonatal mortality remains high at nearly 40 deaths per 1,000 liveborn singletons, despite near-universal skilled attendance and increasing caesarean use. Under current conditions, neither skilled birth attendance nor caesarean delivery, considered in isolation, is associated with large, measurable reductions in early neonatal death; rather, these interventions are embedded in a system in which the highest-risk pregnancies arrive late and where critical elements of intrapartum and neonatal care are unevenly available. Closing the gap to global neonatal survival targets will require moving beyond coverage indicators to ensure that poor and rural women can reach timely, high-quality obstetric and newborn care, and that facilities are equipped to deliver coherent bundles of skilled attendance, safe surgery, and effective neonatal support. If those system-level reforms can be achieved, the modest absolute effects observed here could translate into substantial national reductions in preventable early neonatal deaths.

## Data Availability

The study used only openly available, de-identified human data from the 2018 Nigeria Demographic and Health Survey (NDHS), which were publicly accessible before the initiation of this study. The NDHS microdata (births recode file) can be located and requested free of charge from The Demographic and Health Surveys (DHS) Program website by registering for access and specifying the survey Nigeria DHS 2018 (implemented by the National Population Commission and ICF). Data download pages are available at:
General DHS datasets portal (survey selection and request): https://dhsprogram.com/data/available-datasets.cfm

https://github.com/drsunday-ade/intrapartum-care-neonatal-survival-nigeria-dhs

https://doi.org/10.5281/zenodo.17705787

https://dhsprogram.com/data/available-datasets.cfm

## Abbreviations

ANC: (antenatal care)
BCG: (bacillus Calmette–Guérin vaccine)
CI: (confidence interval)
DAG: (directed acyclic graph)
DHS: (Demographic and Health Surveys)
ICF: (ICF International)
IGME: (Inter-agency Group for Child Mortality Estimation)
IQR: (interquartile range)
MD: (Doctor of Medicine)
NDHS: (Nigeria Demographic and Health Survey)
NPC: (National Population Commission)
OW: (overlap weighting)
PS: (propensity score)
RD: (risk difference)
SBA: (skilled birth attendance)
SD: (standard deviation)
SMD: (standardized mean difference)
STROBE: (Strengthening the Reporting of Observational Studies in Epidemiology)
UN: (United Nations)
UNICEF: (United Nations Children’s Fund)
WHO: (World Health Organization).

## Data and Code Availability

This study used the 2018 Nigeria Demographic and Health Survey (DHS) Births Recode file, accessed under a DHS Program data-use agreement. Individual-level microdata cannot be shared by the authors, but can be requested directly from the DHS Program (https://www.dhsprogram.com). All analysis scripts and non-identifiable outputs are available at GitHub (https://github.com/drsunday-ade/intrapartum-care-neonatal-survival-nigeria-dhs) and archived on Zenodo (https://doi.org/10.5281/zenodo.17705787).

## Ethics Approval and Consent to Participate

The Nigeria 2018 DHS received ethical approval from the National Health Research Ethics Committee of Nigeria and the ICF Institutional Review Board, with informed consent obtained from all participants. This secondary analysis of de-identified public-use data was deemed exempt from additional institutional review, and no further consent was required.

## Authors’ Contributions

S.A.A. conceived the study, designed the analysis, curated the data, ran all statistical analyses, and drafted the manuscript. B.A. contributed to interpretation of findings and critically revised the manuscript for important intellectual content. Both authors approved the final version and agree to be accountable for all aspects of the work.

## Acknowledgments

We thank the DHS Program and the National Population Commission of Nigeria for providing access to the 2018 Nigeria DHS data, and the survey participants whose information made this analysis possible. The views expressed are those of the authors alone.

## Appendix/Supplementary

**Figure 5A.**
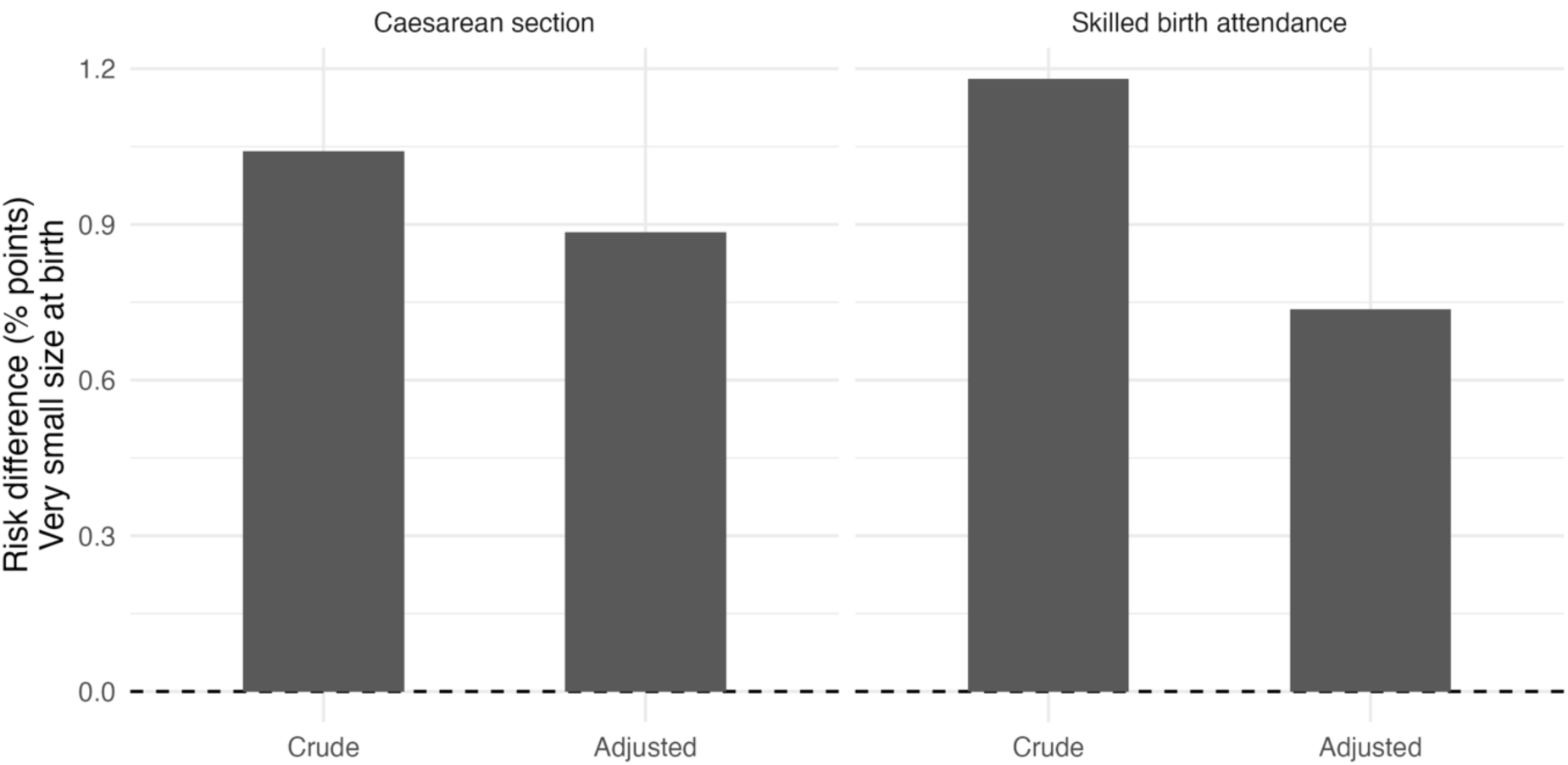
*Crude and overlap-weighted risk differences for “very small” size at birth under skilled birth attendance and caesarean delivery among facility-based singleton births, Nigeria DHS 2018.* Bars show percentage-point risk differences comparing births with versus without skilled attendance (left) and caesarean versus vaginal delivery (right). Error bars represent 95% confidence intervals accounting for the complex survey design and propensity-score overlap weighting.

**Figure 5B.**
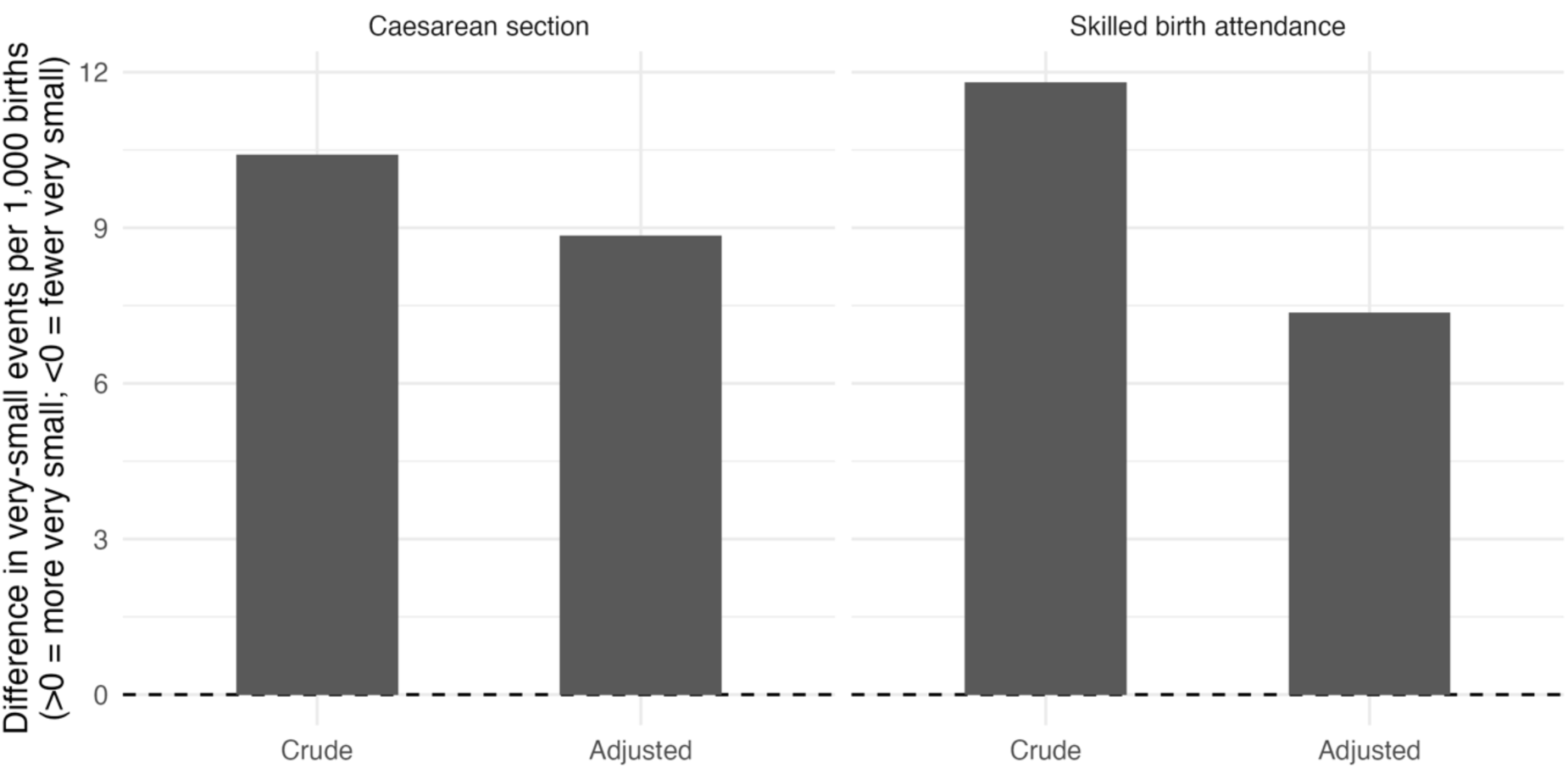
(Supplementary Appendix). *Change in “very small” size-at-birth events per 1,000 facility singleton births under skilled birth attendance and caesarean delivery (crude vs overlap-weighted).* Bars show the absolute difference in the number of very-small births per 1,000 facility singleton births under each exposure contrast, before and after propensity-score overlap weighting; negative values indicate fewer events under the exposed care pattern.

**Supplementary Figure S6B.**
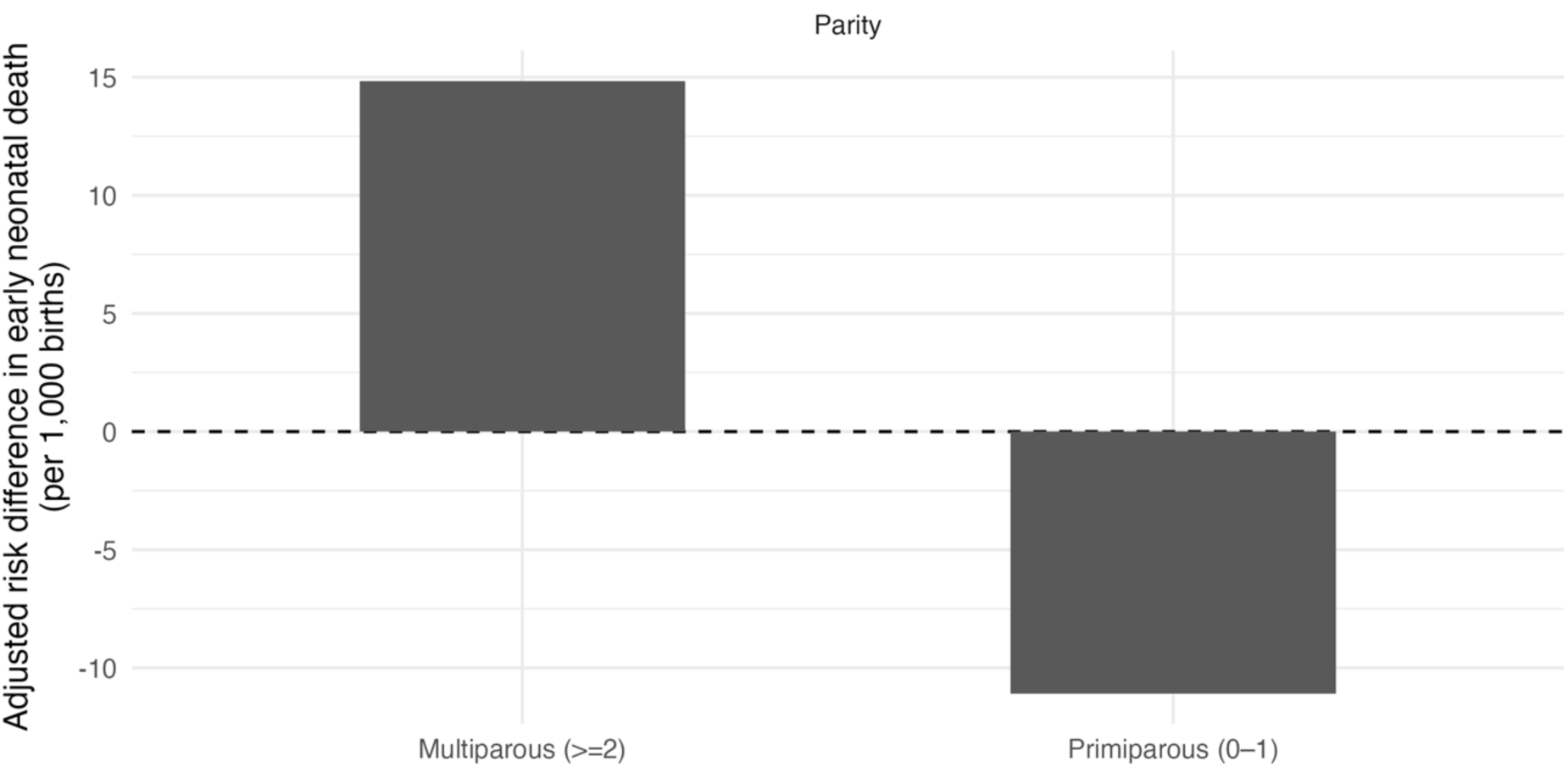
(parity effect modification – C-section). *Overlap-weighted risk difference in early neonatal death per 1,000 births comparing caesarean versus vaginal delivery, by parity group (primiparous vs multiparous), among facility-based singleton births in Nigeria DHS 2018*. Bars show adjusted risk differences on the absolute scale; negative values favour caesarean delivery.

**Supplementary Figure S6C.**
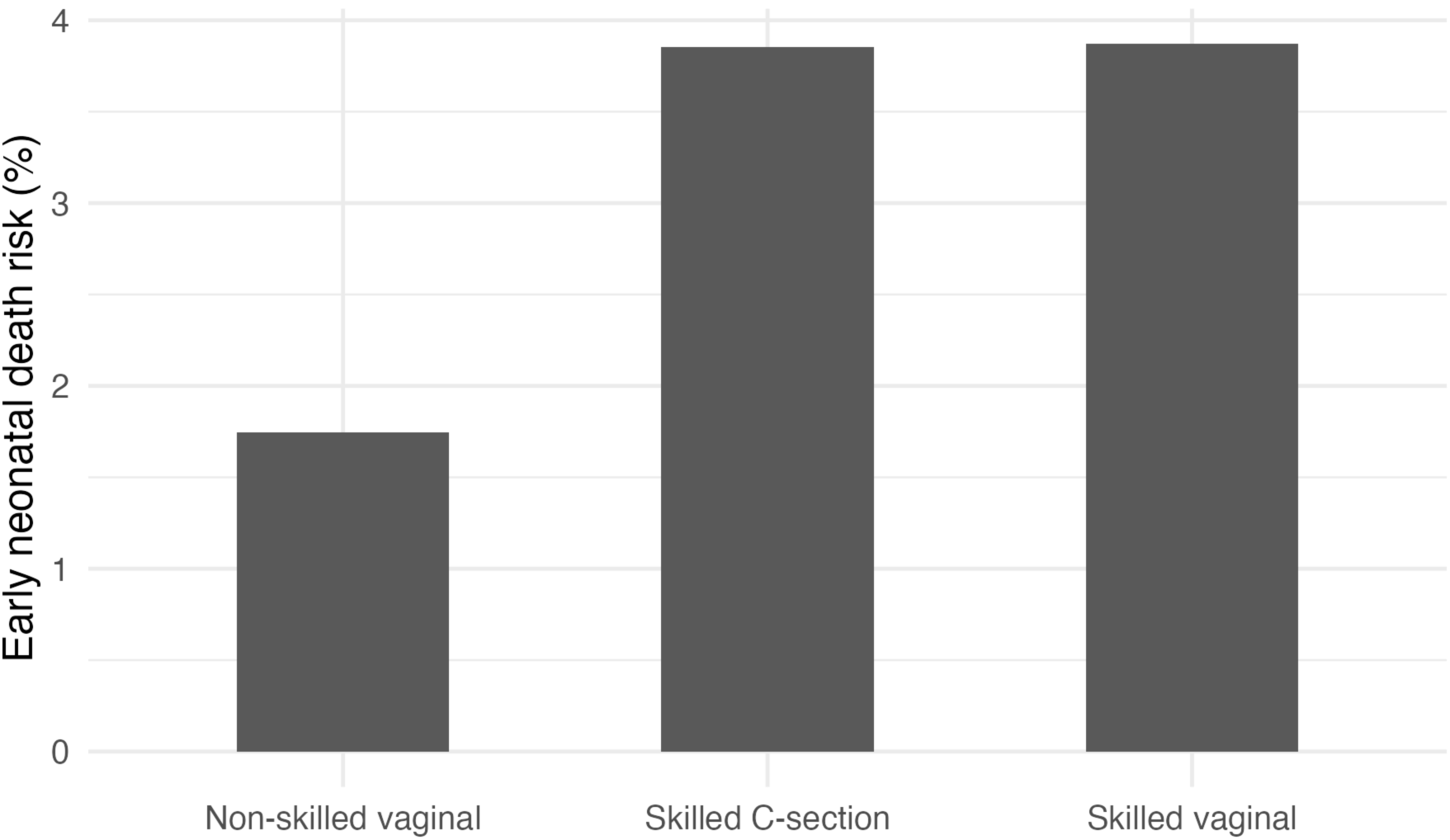
(parity effect modification – SBA). *Overlap-weighted risk difference in early neonatal death per 1,000 births comparing skilled versus non-skilled attendance, by parity group, among facility-based singleton births in Nigeria DHS 2018*. Bars show adjusted absolute differences; error bars represent 95% confidence intervals.

**Supplementary Figure S1.**
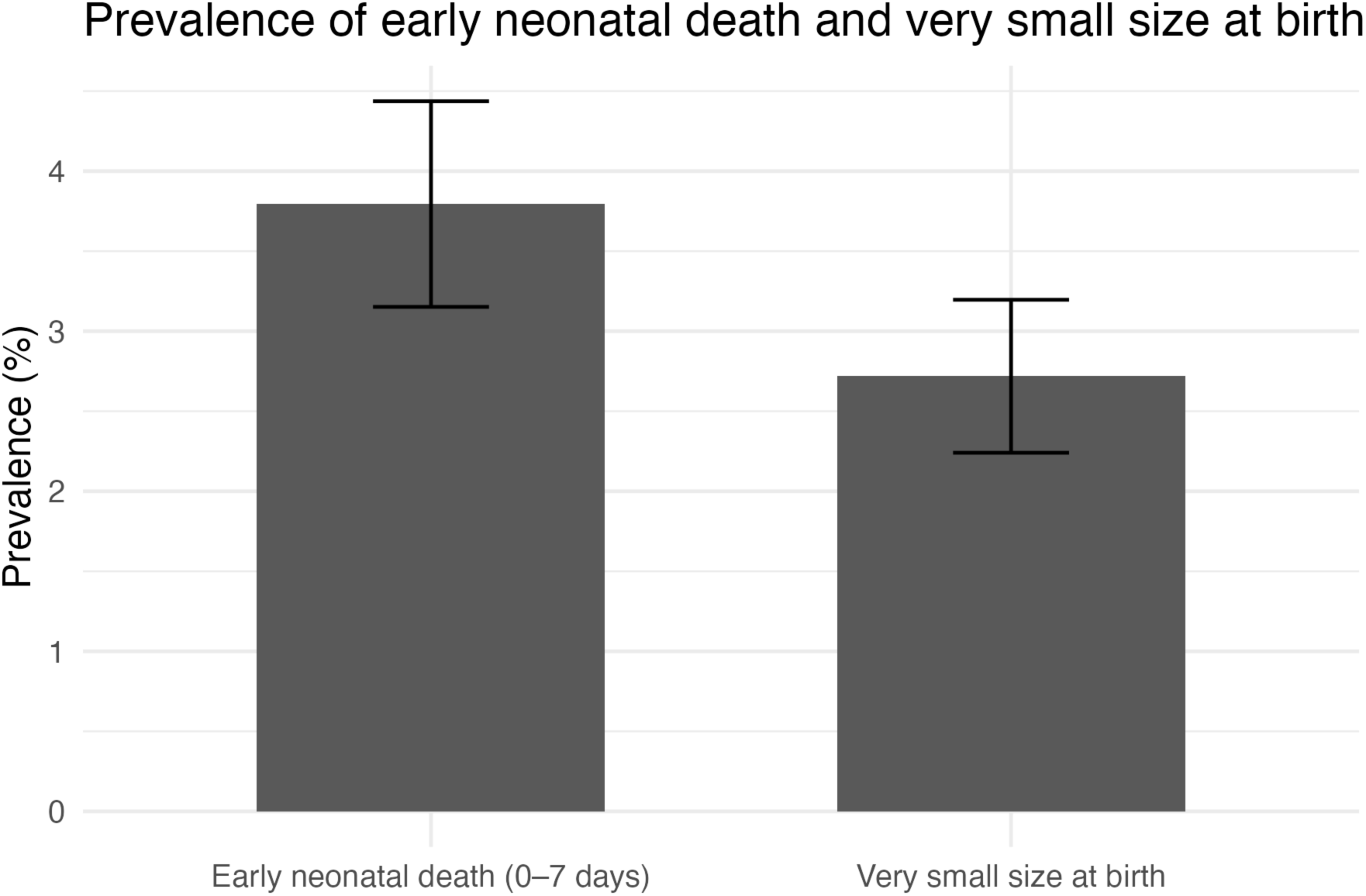
*Prevalence of early neonatal death and “very small” size at birth among facility-based singleton births, Nigeria DHS 2018*. Bars show survey-weighted prevalence with 95% confidence intervals.

**Figure 2A (Supplementary).**
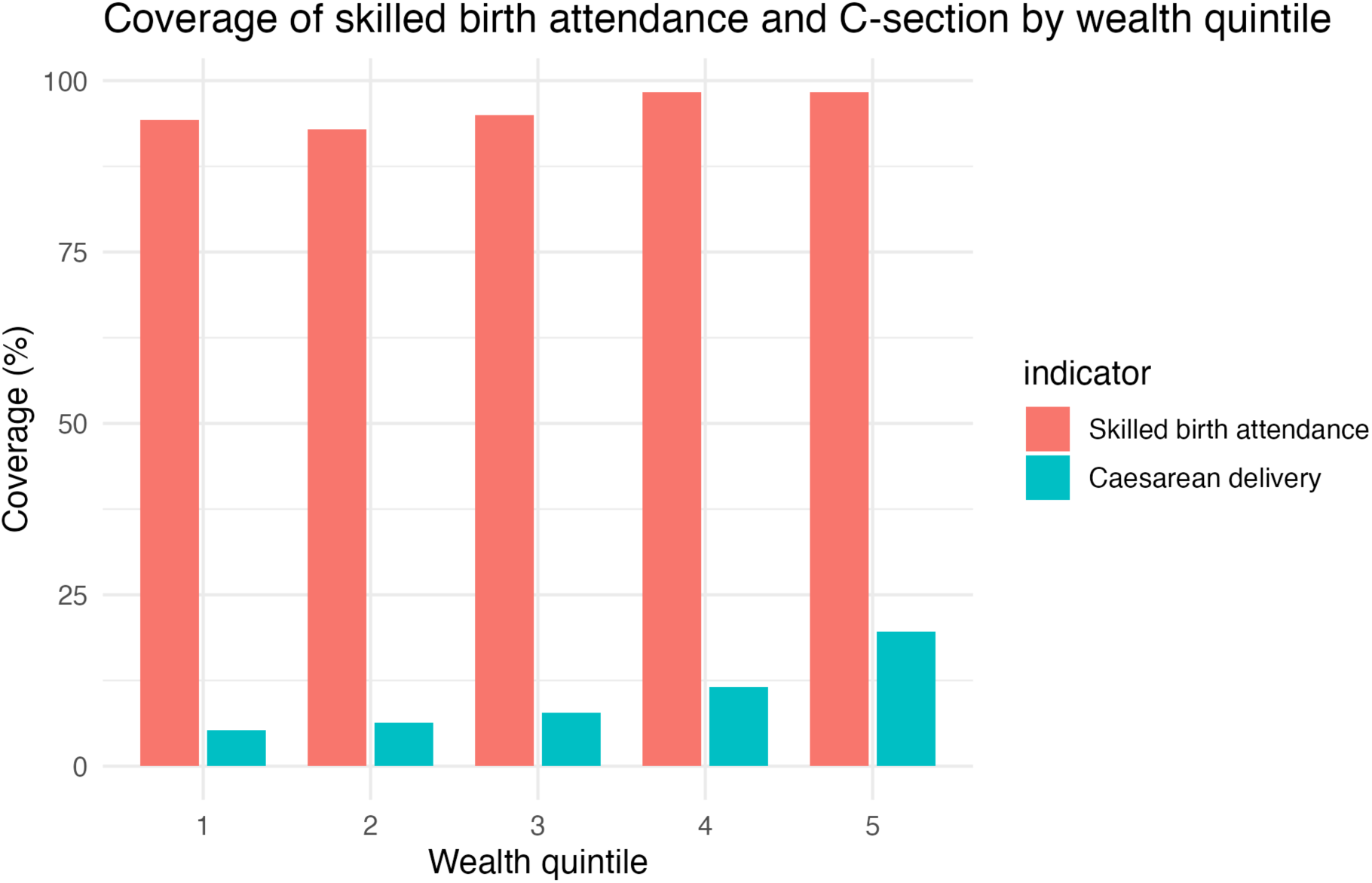
*Coverage of skilled birth attendance and caesarean delivery across wealth quintiles among facility-based singleton births, Nigeria DHS 2018.* Line (or bar) plots show survey-weighted coverage of skilled attendance and caesarean section in each household wealth quintile.

**Supplementary Table S1.** Numeric baseline characteristics among analytic facility singleton births (survey-weighted)

| Characteristic | Weighted mean<br>(SD) | Unweighted<br>n | Weighted<br>births |
| --- | --- | --- | --- |
| Antenatal care visits (number) | 4.8 (3.1) | 5,860 | 5,718.0 |
| Birth order (index within analytic cohort) | 2.9 (1.8) | 5,860 | 5,718.0 |
| Maternal age (years) | 29.6 (6.5) | 5,860 | 5,718.0 |
| Parity (number of prior births) | 2.7 (1.9) | 5,860 | 5,718.0 |

*Note:* Values are survey-weighted means and standard deviations among the final analytic cohort of facility-based singleton births. “Weighted births” are the sum of sampling weights and approximate the number of births represented in the population.

**Supplementary Table S2.** Comparison of included vs excluded births: categorical characteristics (survey-weighted)

| Characteristic | Category | Included (%) | Excluded (%) |
| --- | --- | --- | --- |
| Maternal education | No education | 19.3 | 31.4 |
|  | Primary | 10.4 | 11.2 |
|  | Secondary | 48.5 | 42.0 |
|  | Higher | 21.8 | 15.4 |
| Region | North Central | 13.8 | 15.6 |
|  | North East | 16.8 | 21.9 |
|  | North West | 21.9 | 24.0 |
|  | South East | 18.8 | 13.2 |
|  | South South | 13.8 | 11.1 |
|  | South West | 14.9 | 14.2 |
| Residence | Rural | 41.5 | 61.8 |
|  | Urban | 58.5 | 38.2 |
| Household wealth quintile | Poorest | 10.3 | 20.9 |
|  | Poorer | 12.8 | 15.8 |
|  | Middle | 19.2 | 18.1 |
|  | Richer | 25.9 | 21.7 |
|  | Richest | 31.8 | 23.5 |

*Note:* Percentages are survey-weighted and may not sum to exactly 100% due to rounding. “Included” refers to births in the final analytic cohort; “Excluded” refers to births dropped due to missing outcome, exposure, or covariate data.

**Table 6 (Supplementary Appendix).** Caesarean vs vaginal delivery: crude and overlap-weighted associations with “very small” size at birth.

| Model | Risk of very small size at birth (%) – caesarean | Risk of very small size at birth (%) – vaginal | Risk difference (percentage points) | Risk difference 95% CI | Risk ratio | Risk ratio 95% CI | Odds ratio | Odds ratio 95% CI | Difference in events per 1,000 births |
| --- | --- | --- | --- | --- | --- | --- | --- | --- | --- |
| Crude (survey-weighted) | 3.6 | 2.6 | 1.0 | –0.6 to 2.7 | 1.40 | 0.87 to 2.26 | 1.40 | 0.86 to 2.27 | 10.3 |
| Adjusted (PS overlap-weighted + survey) | 3.5 | 2.7 | 0.9 | –0.8 to 2.5 | 1.35 | 0.80 to 2.28 | 1.35 | 0.80 to 2.29 | 8.6 |

*Note:* “Very small” size at birth is based on the maternal report of size at birth. Overlap-weighted models use propensity-score overlap weights to balance maternal age, parity, education, wealth quintile, residence, region, antenatal care, birth year/order, infant sex, and facility sector.

**Supplementary Table S7.**
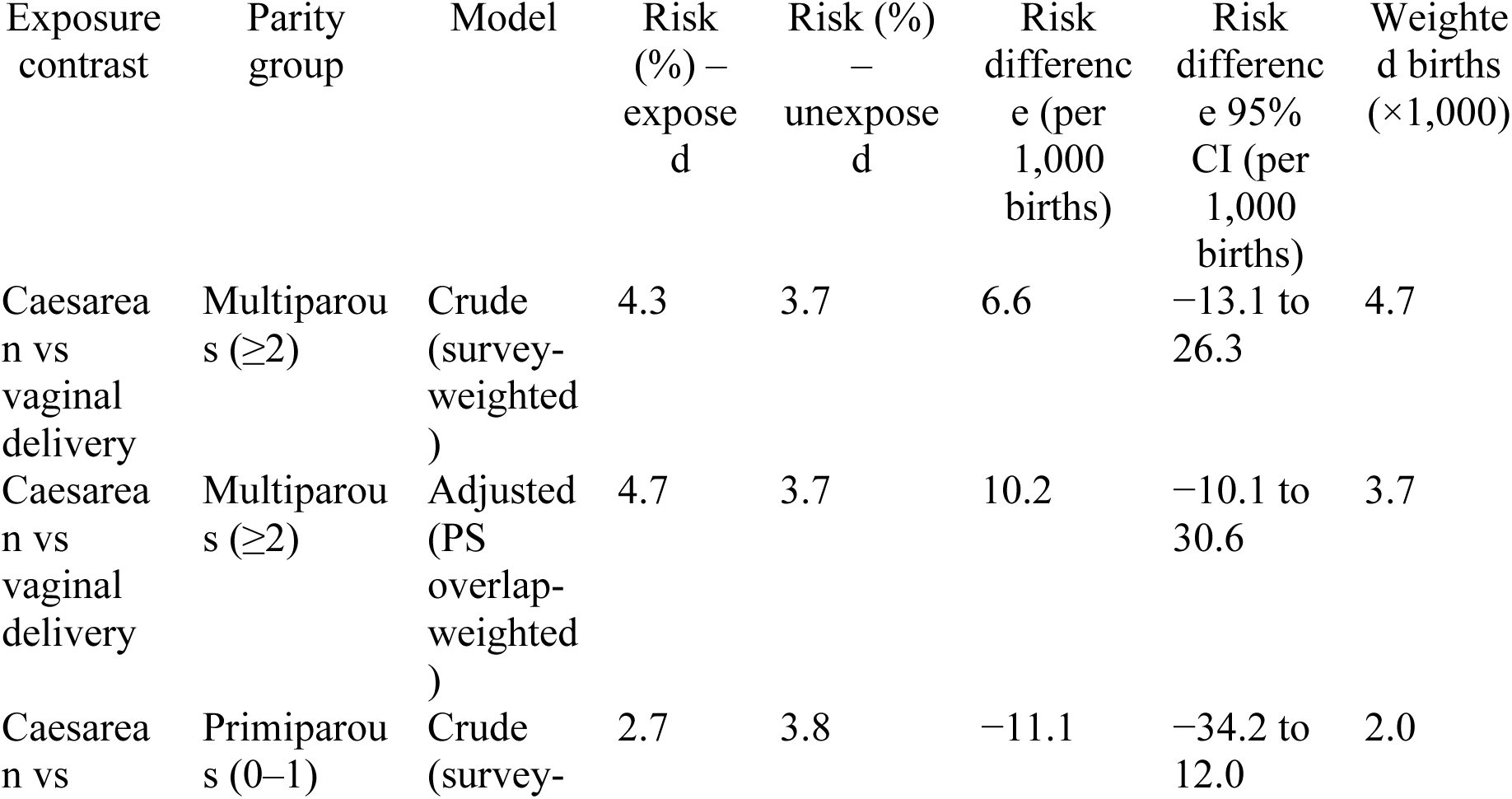
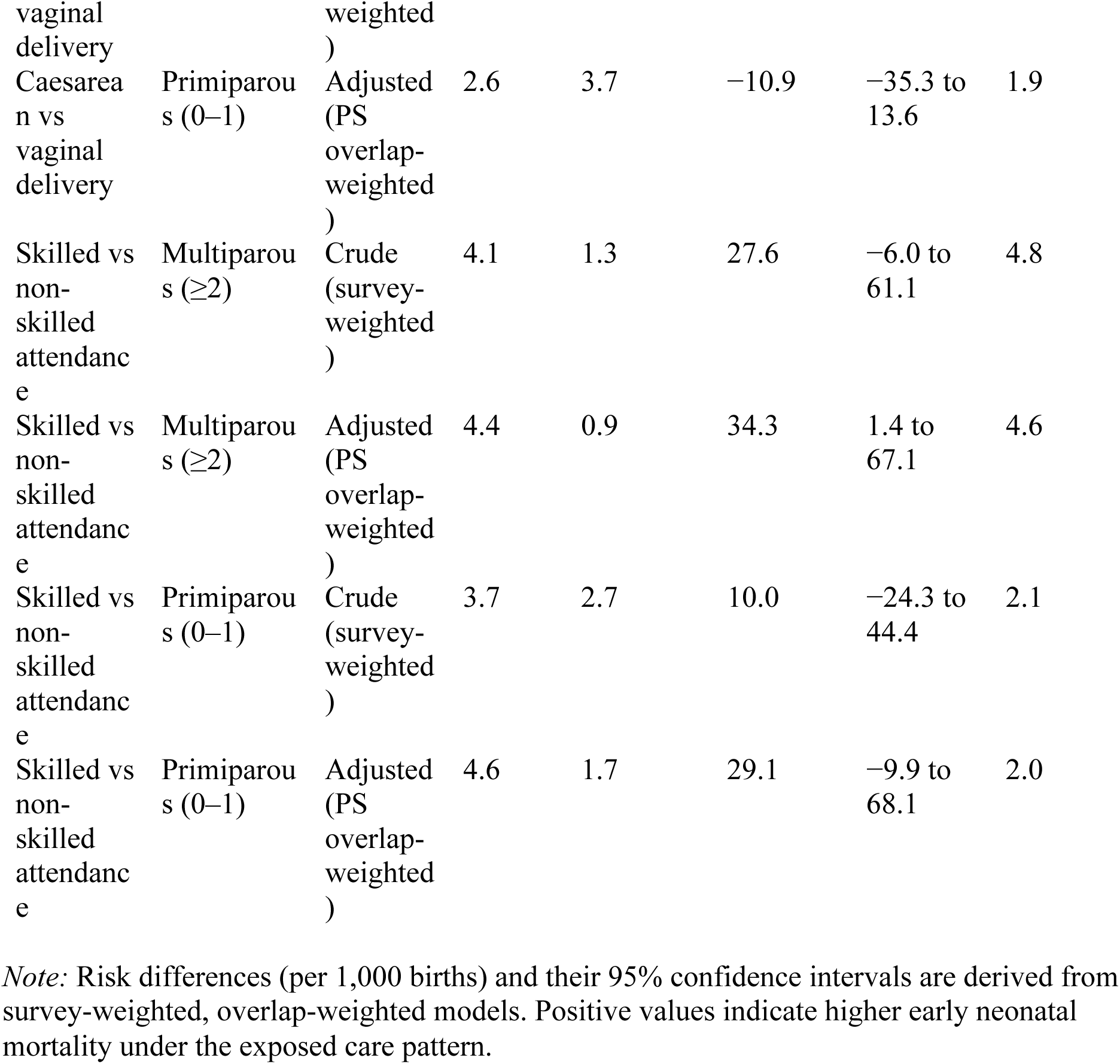
Overlap-weighted effect estimates for caesarean section and skilled attendance, stratified by parity.

| Exposure contrast | Parity group | Model | Risk (%) – exposed | Risk (%) – unexposed | Risk difference (per 1,000 births) | Risk difference 95% CI (per 1,000 births) | Weighted births (×1,000) |
| --- | --- | --- | --- | --- | --- | --- | --- |
| Caesarean vs vaginal delivery | Multiparous (≥2) | Crude (survey-weighted) | 4.3 | 3.7 | 6.6 | –13.1 to 26.3 | 4.7 |
| Caesarean vs vaginal delivery | Multiparous (≥2) | Adjusted (PS overlap-weighted) | 4.7 | 3.7 | 10.2 | –10.1 to 30.6 | 3.7 |
| Caesarean vs vaginal delivery | Primiparous (0–1) | Crude (survey-weighted) | 2.7 | 3.8 | –11.1 | –34.2 to 12.0 | 2.0 |

|  |  |  |  |  |  |  |  |  |
| --- | --- | --- | --- | --- | --- | --- | --- | --- |
| vaginal delivery |  | weighted ) |  |  |  |  |  |  |
| Caesarean vs vaginal delivery | Primiparous (0–1) | Adjusted (PS overlap-weighted ) | 2.6 | 3.7 | –10.9 | –35.3 to 13.6 | 1.9 |  |
| Skilled vs non-skilled attendance | Multiparous ($\geq 2$) | Crude (survey-weighted ) | 4.1 | 1.3 | 27.6 | –6.0 to 61.1 | 4.8 | |
| Skilled vs non-skilled attendance | Multiparous ($\geq 2$) | Adjusted (PS overlap-weighted ) | 4.4 | 0.9 | 34.3 | 1.4 to 67.1 | 4.6 | |
| Skilled vs non-skilled attendance | Primiparous (0–1) | Crude (survey-weighted ) | 3.7 | 2.7 | 10.0 | –24.3 to 44.4 | 2.1 |  |
| Skilled vs non-skilled attendance | Primiparous (0–1) | Adjusted (PS overlap-weighted ) | 4.6 | 1.7 | 29.1 | –9.9 to 68.1 | 2.0 |  |

*Note:* Risk differences (per 1,000 births) and their 95% confidence intervals are derived from survey-weighted, overlap-weighted models. Positive values indicate higher early neonatal mortality under the exposed care pattern.

**Supplementary Table S8.**
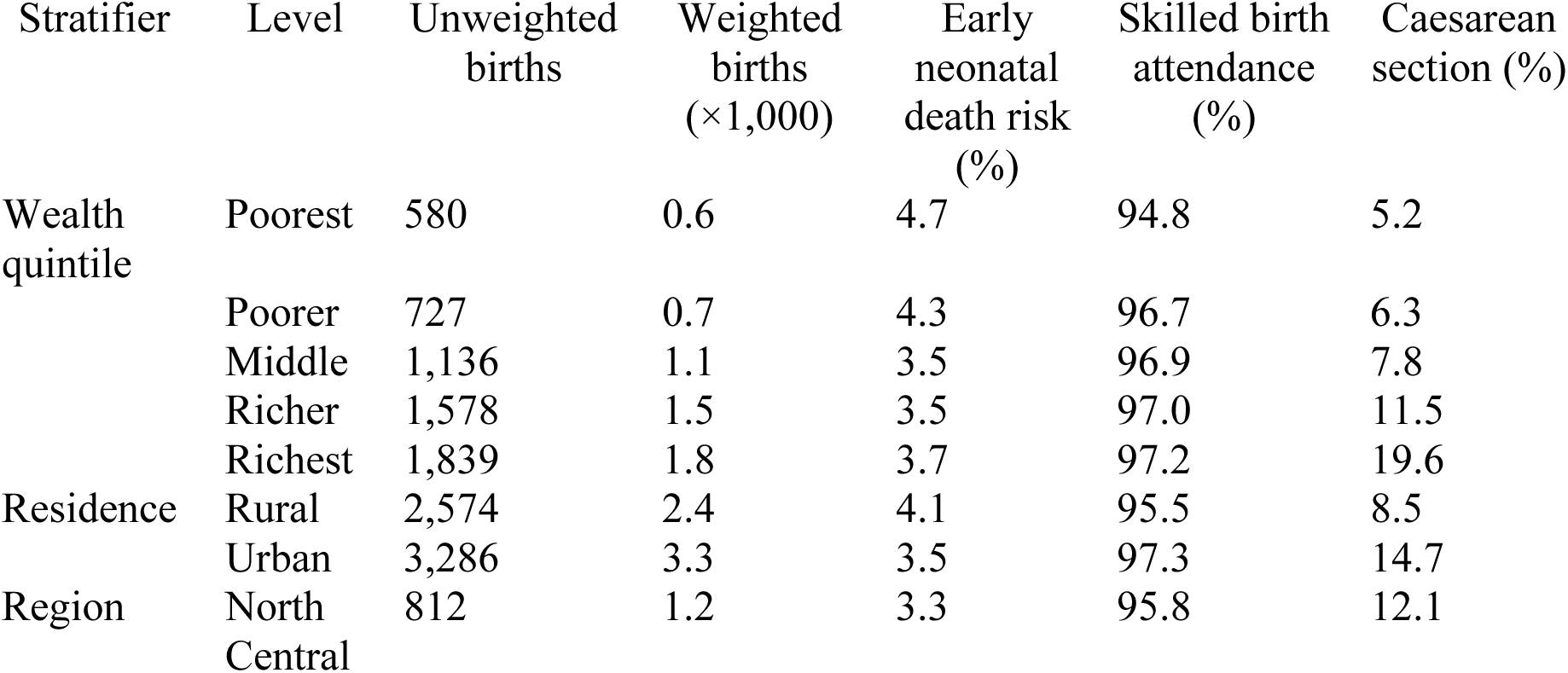
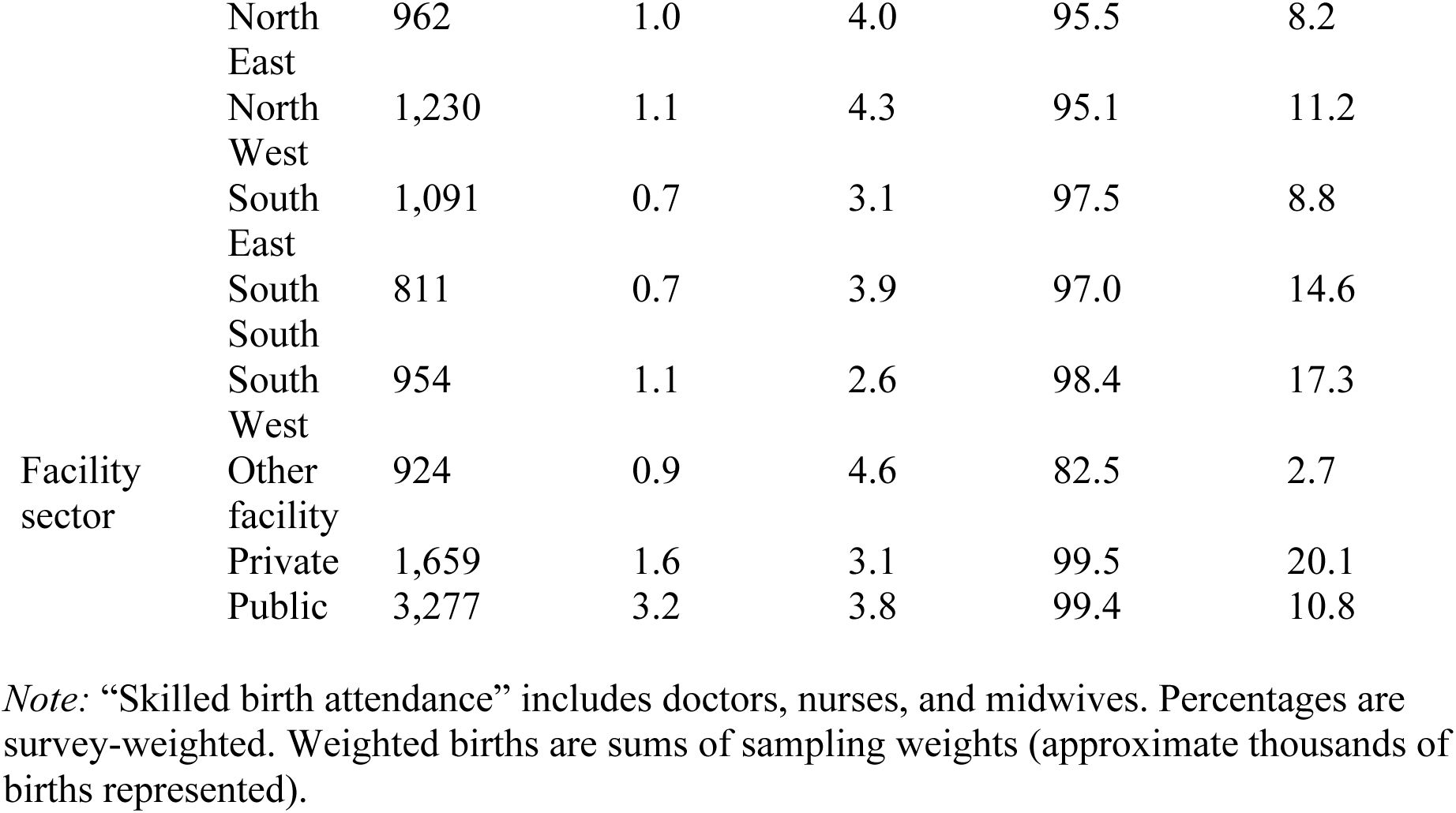
Coverage of skilled birth attendance and caesarean section, and early neonatal mortality by equity strata.

**Supplementary Table S9.** Early neonatal death risk by joint intrapartum care pattern.

| Joint care pattern | Unweighted births | Weighted births | Early neonatal death risk (%) | 95% CI for risk (%) |
| --- | --- | --- | --- | --- |
| Non-skilled vaginal | 201 | 190.4 | 1.7 | −0.1 to 3.5 |
| Skilled caesarean | 702 | 690.2 | 3.8 | 2.5 to 5.0 |
| Skilled vaginal | 4,957 | 4,838.1 | 3.8 | 3.2 to 4.5 |

*Note:* Risks and 95% confidence intervals are survey-weighted. The lower bound for the non-skilled vaginal group slightly below 0% reflects sampling variability and should be interpreted as compatible with very low, but non-negative, risk.

